# Protein-truncating and rare missense variants in *ATM* and *CHEK2* and associations with cancer in UK Biobank whole-exome sequenced data

**DOI:** 10.1101/2024.07.01.24309756

**Authors:** Toqir K. Mukhtar, Naomi Wilcox, Joe Dennis, Xin Yang, Marc Naven, Nasim Mavaddat, John R. B. Perry, Eugene J. Gardner, Douglas F. Easton

**Affiliations:** Centre for Cancer Genetic Epidemiology, Department of Public Health and Primary Care, Strangeways Research Laboratory, University of Cambridge, Cambridge, UK; Department of Primary Care and Public Health, Imperial College London, London, UK; MRC Epidemiology Unit, Institute of Metabolic Science, University of Cambridge, Cambridge, UK; Metabolic Research Laboratory, Institute of Metabolic Science, University of Cambridge, Cambridge, UK; Centre for Cancer Genetic Epidemiology, Department of Oncology, University of Cambridge, Cambridge, UK

**Author notes:** Corresponding author: Toqir K. Mukhtar, Centre for Cancer Genetic Epidemiology, Department of Public Health and Primary Care, Strangeways Research Laboratory, Worts’ Causeway, University of Cambridge, Cambridge CB1 8RN, UK.

## Abstract

**Background:** Deleterious germline variants in *ATM* and *CHEK2* have been associated with a moderately increased risk of breast cancer. Risks for other cancers remain unclear, and require further investigation.

**Methods:** Cancer associations for coding variants in *ATM* and *CHEK2* were evaluated using whole-exome sequenced data from UK Biobank linked to cancer registration data (348,488 participants), and analysed both as a retrospective case-control and a prospective cohort study. Odds ratios, hazard ratios, and combined relative risks (RRs) were estimated by cancer type and gene. Separate analyses were performed for protein-truncating variants (PTVs) and rare missense variants (rMSVs; allele frequency <0·1%).

**Results:** PTVs in *ATM* were associated with increased risks of nine cancers at p<0·001 (pancreas, oesophagus, lung, melanoma, breast, ovary, prostate, bladder, lymphoid leukaemia [LL]), and two at p<0·05 (colon, diffuse non-Hodgkin’s lymphoma [DNHL]). Carriers of rMSVs had increased risks of four cancers (p<0·05: stomach, pancreas, prostate, Hodgkin’s disease [HD]). RRs were highest for breast, prostate, and any cancer where rMSVs lay in the FAT or PIK domains, and had a CADD score in the highest quintile.

PTVs in *CHEK2* were associated with three cancers at p<0·001 (breast, prostate, HD), and six at p<0·05 (oesophagus, melanoma, ovary, kidney, DNHL, myeloid leukaemia). Carriers of rMSVs had increased risks of five cancers (p<0·001: breast, prostate, LL; p<0·05: melanoma, multiple myeloma).

**Conclusion:** PTVs in *ATM* and *CHEK2* are associated with a wide range of cancers, with the highest RR for pancreatic cancer in *ATM* PTV carriers. These findings can inform genetic counselling of carriers.

**WHAT IS ALREADY KNOWN ON THIS TOPIC:**

- While previous research shows there is evidence for association between variants in *ATM* or *CHEK2* and multiple cancer types in individual smaller studies, the associations have not been consistently evaluated across all cancer types and, with the exception of breast cancer, the strengths of association are unclear.

**WHAT THIS STUDY ADDS:**

- We examined data from a large cohort study to derive relative and absolute risks for all cancer types for carriers of PTVs and rMSVs in *CHEK2* and *ATM* .
- *ATM* PTVs were associated with significantly increased risk for 11 of 23 sites examined (nine at p<0·001), with the relative risk being highest for pancreatic cancer (approximately seven-fold). Carriers of rMSVs had increased risks of four cancers, with a RR of approximately 1·5.
- For *CHEK2* PTVs, statistically significant risks were observed for seven of the 21 sites examined (one at p<0·001). Carriers of rMSVs had increased risks of five cancers with the risk being highest for lymphoid leukaemia (approximately two-fold).

**HOW THIS STUDY MIGHT AFFECT RESEARCH, PRACTICE OR POLICY:**

- *ATM* and *CHEK2* are included on many cancer gene panels used in family cancer clinics, and the risk estimates from these analyses can inform genetic counselling for carriers.
- The estimated absolute risks for pancreatic cancer in *ATM* PTV carriers (11% in males and 8% in females by age 85) are notably higher than for other major pancreatic susceptibility genes including BRCA2, CDK2NA, and PALB2. Our findings can also inform NICE guidelines for pancreatic cancer, which do not currently include *ATM* .

## Introduction

Gene panel testing for cancer susceptibility is now an important part of clinical practice. To provide reliable genetic counselling, it is important to be able to provide risks for all cancer types. Many susceptibility genes, notably *BRCA1, BRCA2*, and the mismatch repair genes, are associated with multiple types of cancer (1, 2), but in general, cancer pleiotropy is poorly understood. Here we consider rare variants in two genes, *CHEK2* and *ATM*, that are included on many cancer gene panels. Variants in these genes are known to be associated with moderately increased risks of breast cancer, but the risks of many other cancer types are unclear.

*CHEK2* (*Checkpoint Kinase 2*) encodes a protein kinase, activated in response to DNA damage (3), and with a role in cell-cycle arrest, DNA repair, and cell death (4, 5). The most common protein-truncating variant (PTV) in Western European populations is the *CHEK2*\*1100delC. Previous research has shown an association between the 1100delC deletion and risk of breast cancer in women, with the estimated risk being approximately two-fold (6, 7). A similar association has been seen for other PTVs in aggregate (6, 8). Rare *CHEK2* MSVs, in aggregate, have also been associated with breast cancer risk, with a RR of ∼1.4 (6). There is evidence that the breast cancer risk differs by variant, using *in-silico* and functional classifications of deleteriousness (9).

Previous studies have found some evidence of an association between PTVs and cancer other than breast cancer, with most evidence being based on the 1100delC variant: these include increased risks of prostate cancer (10); colorectal cancer (11, 12); malignant melanoma, gastric cancer, and bladder cancer (13). Previous studies have reported evidence that carriers of the I157T variant have increased risk of prostate (10), colorectal (14), ovarian (15), pancreatic (16), gastric (17), thyroid (18), kidney (19), and colon cancer (19). There is some evidence of a decreased risk of lung cancer (20). *CHEK2* PTVs have also been shown to influence ovarian ageing in women through inhibited DNA damage sensing (21).

*ATM* (*Ataxia-Telangiectasia Mutated*) (22) encodes a protein kinase involved in DNA repair, apoptosis, and regulation of cell-cycle checkpoints (22, 23). It is recruited and activated by double-strand DNA breaks. *ATM* activates *CHEK2* through phosphorylation. Deleterious mutations in *ATM* cause the rare autosomal recessive disorder Ataxia Telangiectasia (A-T). A-T patients have a large excess risk of cancers in childhood, in particular, lymphomas and leukaemias, but are also at increased risk of adult cancers, including breast cancer. There is evidence both from studies of families of A-T patients (24), and from large case-control studies, of an association between pathogenic variants in *ATM* and increased risk of breast cancer (6), with an estimated RR of approximately twofold. Previous studies indicate subsets of rMSVs from the PIK, FAT, and FATC domains are also associated with an excess risk of breast cancer (25).

For cancers other than breast, there is evidence for an association between mutations in *ATM* and cancer of the prostate (26), and pancreas (27). Some studies have found evidence for a higher risk of colorectal cancer (24), melanoma (28), gastric, ovarian, and lung cancer (29, 30).

While there is evidence for association between variants in *ATM* or *CHEK2* and multiple cancer types, the associations have not been consistently evaluated across all cancer types and, with the exception of breast cancer, the strengths of association are unclear. In this paper, we examine data from a large cohort study to derive relative and absolute risks for all cancer types, for carriers of PTVs and rMSVs in *CHEK2* and *ATM*. We also examine the evidence for variation in risk for rMSVs according to previously defined *in-silico* classifications.

## Materials and methods

### Dataset

UK Biobank is a population-based, prospective study of approximately 500,000 participants in the United Kingdom (31). Participants were enrolled between 2006 and 2010 and were aged between 40 and 69 on recruitment. Whole-exome sequence (WES) data were available at the time of analysis for 454,756 UK Biobank participants (32). Details of the WES variant calling, filtering, and classification are given in Supplemental Material. In *ATM*, rMSVs were categorised by functional protein domain (33) and pathogenicity score predicted by the Combined Annotation Dependent Depletion algorithm (CADD; version 1.6) (34). In CHEK2, rMSVs were classified as high or low-risk using the deleteriousness score given by the Helix algorithm (version; 4.4.1) (35). These classifications were used as they provided the best discrimination for breast cancer risk in the analysis of the large BRIDGES dataset.

Cancers occurring in study participants were identified through linkage to the national cancer registry data for England, Wales, and Scotland (36). Invasive cancers (codes C00-C97) and breast carcinoma-in-situ (CIS; code D05), defined using the 10th revision of the International Classification of Diseases (37), were included as endpoints. Self-reported cancers were not included.

### Study design

UK Biobank includes both a retrospective (based on data at study entry) and a prospective component (Figure 1). The retrospective analyses were based on cases diagnosed with cancer before entry to the study. Controls were individuals who had neither a self nor registry-reported cancer before entry, other than non-melanoma skin cancer. A total of 348,488 males and females were included, of whom 18,838 were diagnosed with at least one cancer before the day of first assessment.

**Figure 1:**
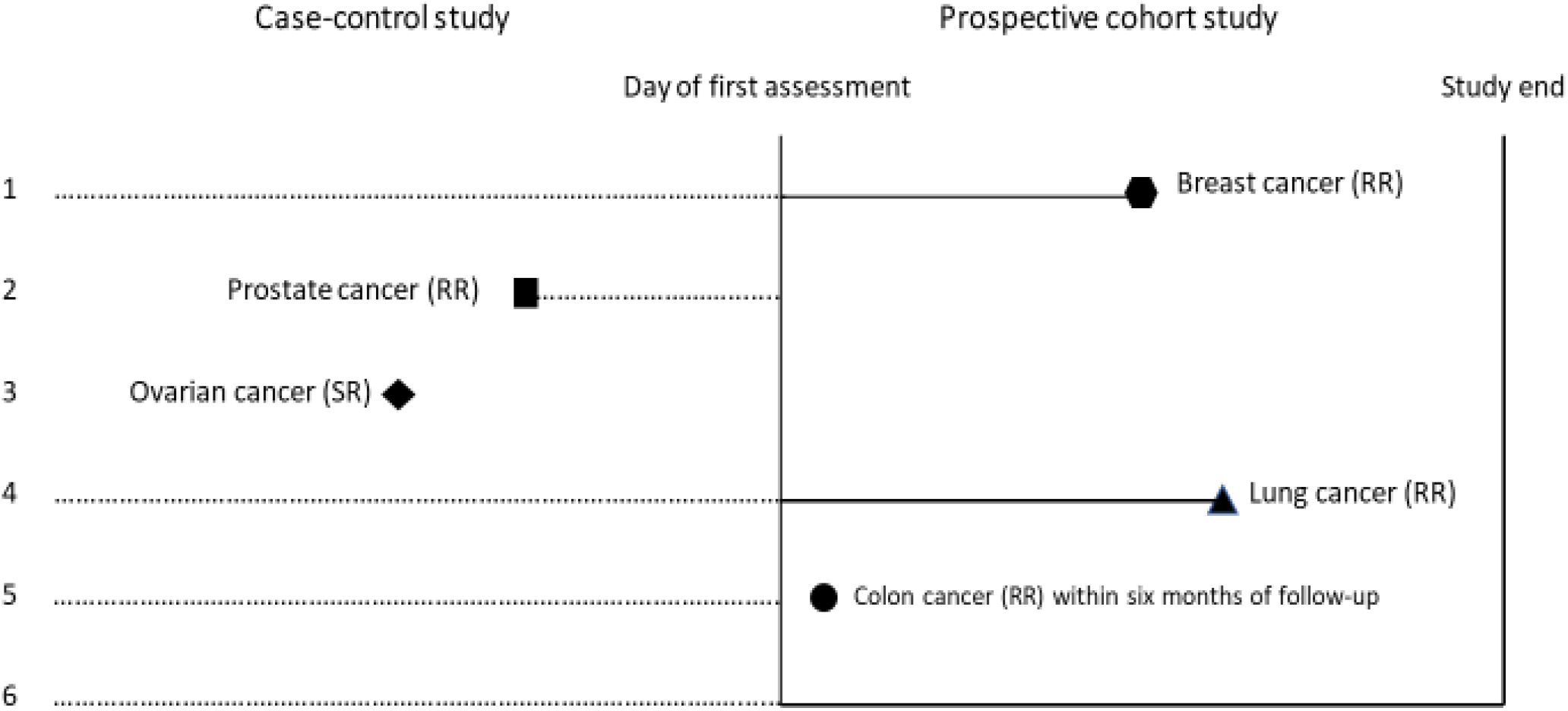
Case-control and prospective cohort studies generatedfrom UK Biobank data (RR) – Registry-Reported cancer (SR) – Self-Reported cancer Day of first assessment: Start of observation for the prospective study. Person 1: Registry-reported breast cancer two years after the day of first assessment, and no prior cancers. Contributes to both studies. Person 2: Registry-reported prostate cancer prior to the first day of assessment. Case-control study, only. Person 3: Self-reported ovarian cancer prior to the first day of assessment, and no registry-reported cancers. Excluded from both studies. Person 4: Registry-reported lung cancer 3 years after the day of first assessment. Contributes to both studies. Person 5: Registry-reported colon cancer 4 months after the start of observation. Case-control study, only. Person 6: No self-reported or registry-reported cancers. Contributes to both studies.

The prospective analysis included individuals without a self-reported cancer or registry reported cancer before the start of follow-up, or a registry-reported cancer within six months of follow-up (except for non-melanoma skin cancer). Thus, follow-up started six months from the day of first assessment for the study, to avoid inadvertent inclusion of cases where onset was prior to entry. The event indicator was first diagnosis of any cancer by type, and follow-up was censored at the earliest of first diagnosed cancer of any type, date of death, or a last follow-up date dependent upon the location of the assessment centre attended by participants (for England this was: December 31, 2020; Scotland: November 30, 2021; and Wales: December 31, 2016). A total of 328,919 individuals were included, with 37,802 cancers diagnosed over 3,484,613 person years of follow-up.

### Statistical analyses

Since most variants were too rare to estimate the associated risk, we conducted burden analyses which evaluated the risks associated with carrying any one of a set of variants: PTVs or rMSVs. rMSVs were further subdivided by functional protein domain and prediction scores.

In the retrospective dataset, logistic regression was used to estimate the odds ratio (OR) for each cancer type associated with carrying either a PTV or rMSV. For the prospective analyses, Cox proportional hazard regression models were used to estimate the hazard ratio (HR) for any cancer, and for each cancer type, associated with carrying either a PTV or rMSV. All models included sex and 10 ancestry informative principal components (35) as covariates. Since the retrospective and prospective components are essentially independent, we also computed overall relative risk estimates combining the retrospective and prospective estimates (See Supplemental Material). We investigated whether the HRs varied by age by fitting HRs by age-group (<50, 50-59, 60-69, 70-79, and ≥80 years) and by fitting models in which the log(HR) varied linearly with age, using the tt() function in R.

Cumulative risks for cancer were estimated by combining HR estimates from the prospective analyses with population cancer incidences in the United Kingdom, 2016-2018 (38), using the same approach as that used by Schmidt et al., (39). Where there was evidence of a trend in the HR by age, time dependent HR risk estimates were used, otherwise a constant HR was assumed.

Binary logistic regression and Cox proportional hazard regression analyses were conducted using R version 4.1.0. Meta-analyses were also conducted in R using the meta package version 6.5-0, and the metagen function. All p-values were two-sided, with p<0·05 considered significant.

## Results

### PTVs in ATM

In the pooled analysis (Table 1), PTVs in *ATM* were associated with increased risks of 11 cancers: oesophagus (RR: 3·90 [95% CI: 1·85 to 8·20, p=3·43 x 10^−4^]); colon (2·18 [1·37 to 3·48, p=1·04 x 10^−3^]); pancreas (7·35 [4·27 to 12·66, p=6·53 x 10^−13^]); lung (2·64 [1·61 to 4·32, p=1·18 x 10^−4^]); melanoma (2·28 [1·44 to 3·59, p=4·18 x 10^−4^]), breast (2·27 [1·77 to 2·91, p=1·18 x 10^−10^]); ovary (3·20 [1·66 to 6·16, p=5·12 x 10^−4^]); prostate (2·35 [1·78 to 3·11, p=2·41 x 10^−9^]); bladder (3·74 [1·94 to 7·21, p=8·30 x 10^−5^]); diffuse non-Hodgkin’s Lymphoma (DNHL) (3·18 [1·51 to 6·69, p=2·28 x 10^−3^]); and lymphoid leukaemia (LL) (4·42 [2·11 to 9·29, p=8·67 x 10^−5^]). PTVs were also associated with breast carcinoma-in-situ (CIS) (3·22 [1·96 to 5·28, p=4·05 x 10^−6^]).

**Table 1:**
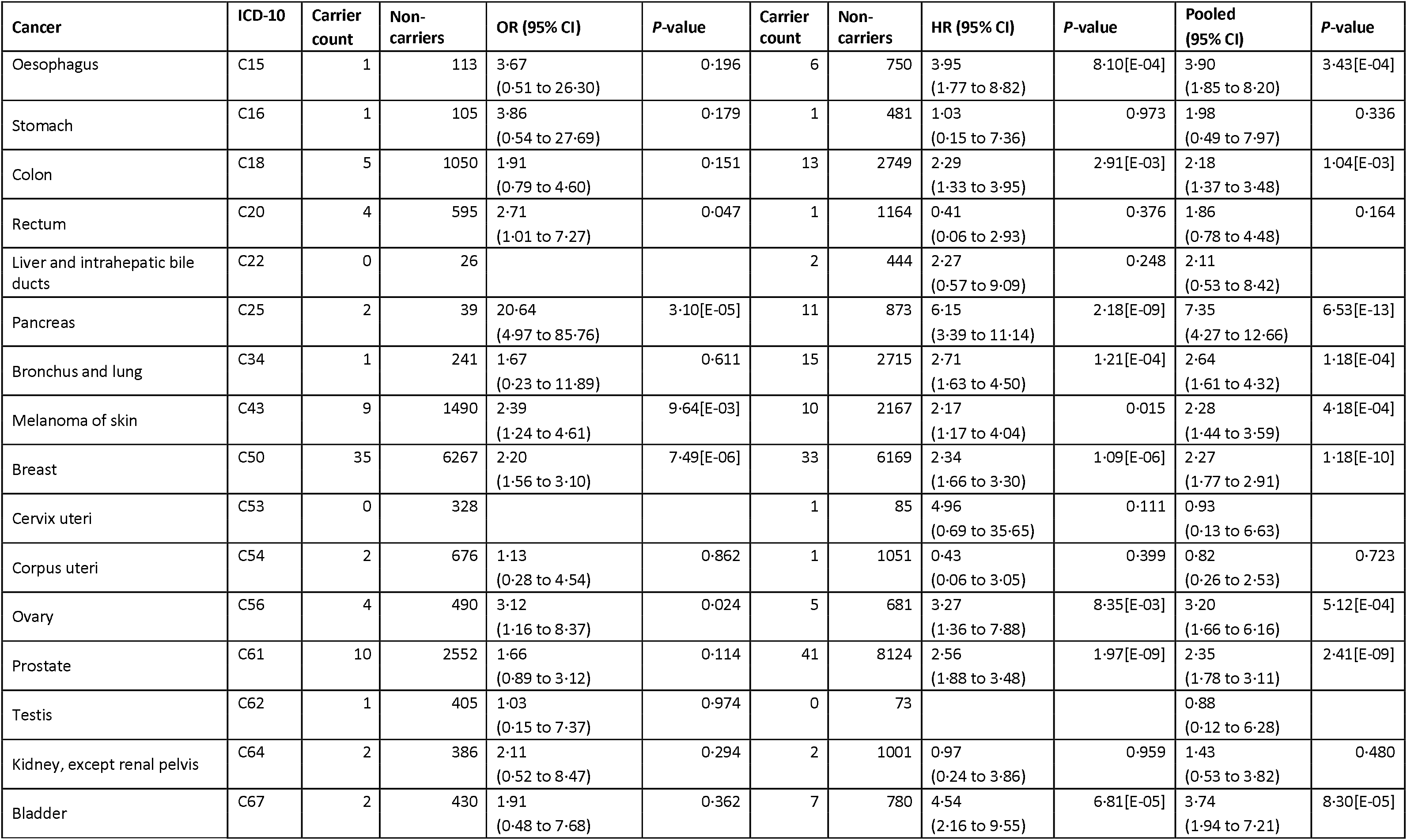
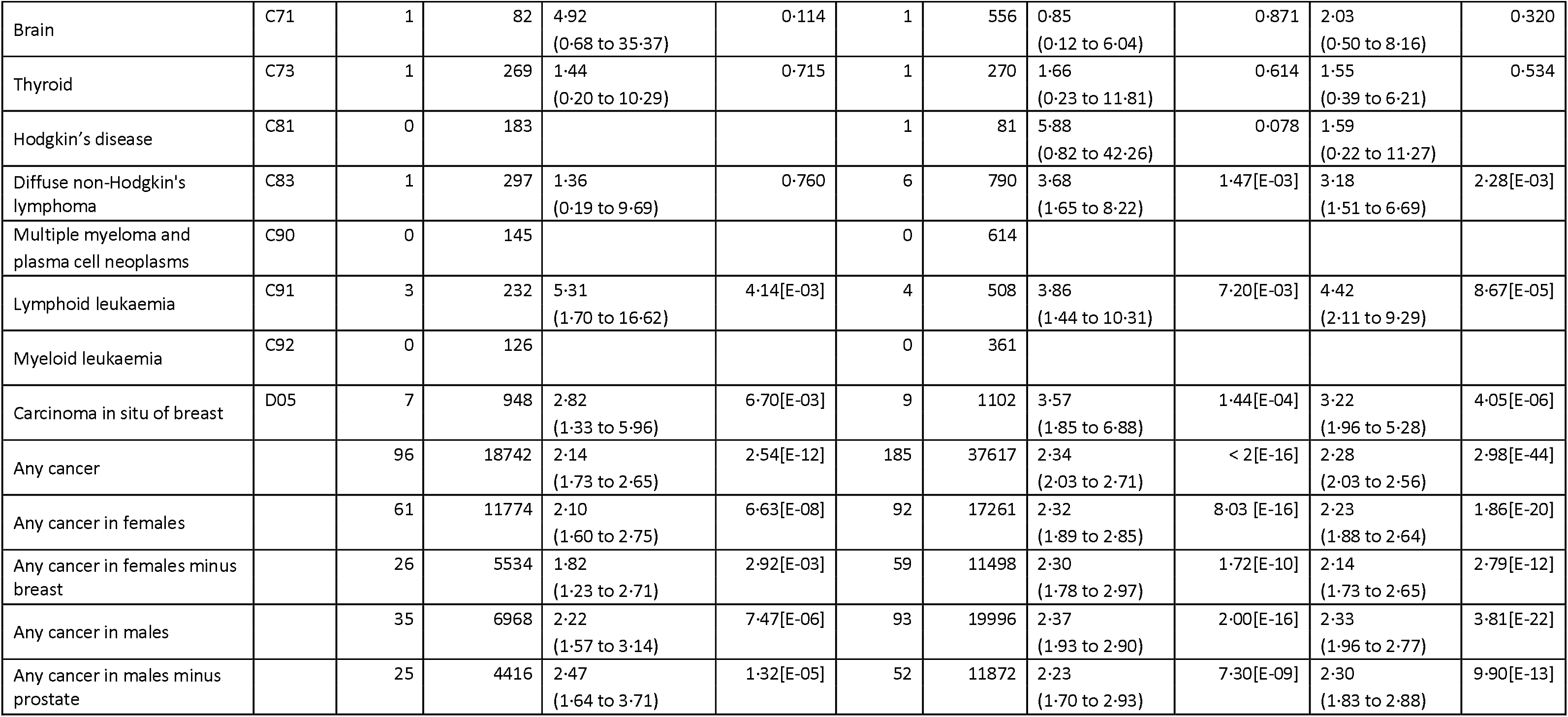
Odds ratios, hazard ratios, and pooled estimates for the association of PTVs in ATM with cancer by type.

| Cancer | ICD-10 | Carrier count | Non-carriers | OR (95% CI) | P-value | Carrier count | Non-carriers | HR (95% CI) | P-value | Pooled (95% CI) | P-value |
| --- | --- | --- | --- | --- | --- | --- | --- | --- | --- | --- | --- |
| Oesophagus | C15 | 1 | 113 | 3.67<br>(0.51 to 26.30) | 0.196 | 6 | 750 | 3.95<br>(1.77 to 8.82) | 8.10[E-04] | 3.90<br>(1.85 to 8.20) | 3.43[E-04] |
| Stomach | C16 | 1 | 105 | 3.86<br>(0.54 to 27.69) | 0.179 | 1 | 481 | 1.03<br>(0.15 to 7.36) | 0.973 | 1.98<br>(0.49 to 7.97) | 0.336 |
| Colon | C18 | 5 | 1050 | 1.91<br>(0.79 to 4.60) | 0.151 | 13 | 2749 | 2.29<br>(1.33 to 3.95) | 2.91[E-03] | 2.18<br>(1.37 to 3.48) | 1.04[E-03] |
| Rectum | C20 | 4 | 595 | 2.71<br>(1.01 to 7.27) | 0.047 | 1 | 1164 | 0.41<br>(0.06 to 2.93) | 0.376 | 1.86<br>(0.78 to 4.48) | 0.164 |
| Liver and intrahepatic bile ducts | C22 | 0 | 26 |  |  | 2 | 444 | 2.27<br>(0.57 to 9.09) | 0.248 | 2.11<br>(0.53 to 8.42) |  |
| Pancreas | C25 | 2 | 39 | 20.64<br>(4.97 to 85.76) | 3.10[E-05] | 11 | 873 | 6.15<br>(3.39 to 11.14) | 2.18[E-09] | 7.35<br>(4.27 to 12.66) | 6.53[E-13] |
| Bronchus and lung | C34 | 1 | 241 | 1.67<br>(0.23 to 11.89) | 0.611 | 15 | 2715 | 2.71<br>(1.63 to 4.50) | 1.21[E-04] | 2.64<br>(1.61 to 4.32) | 1.18[E-04] |
| Melanoma of skin | C43 | 9 | 1490 | 2.39<br>(1.24 to 4.61) | 9.64[E-03] | 10 | 2167 | 2.17<br>(1.17 to 4.04) | 0.015 | 2.28<br>(1.44 to 3.59) | 4.18[E-04] |
| Breast | C50 | 35 | 6267 | 2.20<br>(1.56 to 3.10) | 7.49[E-06] | 33 | 6169 | 2.34<br>(1.66 to 3.30) | 1.09[E-06] | 2.27<br>(1.77 to 2.91) | 1.18[E-10] |
| Cervix uteri | C53 | 0 | 328 |  |  | 1 | 85 | 4.96<br>(0.69 to 35.65) | 0.111 | 0.93<br>(0.13 to 6.63) |  |
| Corpus uteri | C54 | 2 | 676 | 1.13<br>(0.28 to 4.54) | 0.862 | 1 | 1051 | 0.43<br>(0.06 to 3.05) | 0.399 | 0.82<br>(0.26 to 2.53) | 0.723 |
| Ovary | C56 | 4 | 490 | 3.12<br>(1.16 to 8.37) | 0.024 | 5 | 681 | 3.27<br>(1.36 to 7.88) | 8.35[E-03] | 3.20<br>(1.66 to 6.16) | 5.12[E-04] |
| Prostate | C61 | 10 | 2552 | 1.66<br>(0.89 to 3.12) | 0.114 | 41 | 8124 | 2.56<br>(1.88 to 3.48) | 1.97[E-09] | 2.35<br>(1.78 to 3.11) | 2.41[E-09] |
| Testis | C62 | 1 | 405 | 1.03<br>(0.15 to 7.37) | 0.974 | 0 | 73 |  |  | 0.88<br>(0.12 to 6.28) |  |
| Kidney, except renal pelvis | C64 | 2 | 386 | 2.11<br>(0.52 to 8.47) | 0.294 | 2 | 1001 | 0.97<br>(0.24 to 3.86) | 0.959 | 1.43<br>(0.53 to 3.82) | 0.480 |
| Bladder | C67 | 2 | 430 | 1.91<br>(0.48 to 7.68) | 0.362 | 7 | 780 | 4.54<br>(2.16 to 9.55) | 6.81[E-05] | 3.74<br>(1.94 to 7.21) | 8.30[E-05] |
| Brain | C71 | 1 | 82 | 4.92<br>(0.68 to 35.37) | 0.114 | 1 | 556 | 0.85<br>(0.12 to 6.04) | 0.871 | 2.03<br>(0.50 to 8.16) | 0.320 |
| Thyroid | C73 | 1 | 269 | 1.44<br>(0.20 to 10.29) | 0.715 | 1 | 270 | 1.66<br>(0.23 to 11.81) | 0.614 | 1.55<br>(0.39 to 6.21) | 0.534 |
| Hodgkin's disease | C81 | 0 | 183 |  |  | 1 | 81 | 5.88<br>(0.82 to 42.26) | 0.078 | 1.59<br>(0.22 to 11.27) |  |
| Diffuse non-Hodgkin's lymphoma | C83 | 1 | 297 | 1.36<br>(0.19 to 9.69) | 0.760 | 6 | 790 | 3.68<br>(1.65 to 8.22) | 1.47[E-03] | 3.18<br>(1.51 to 6.69) | 2.28[E-03] |
| Multiple myeloma and plasma cell neoplasms | C90 | 0 | 145 |  |  | 0 | 614 |  |  |  |  |
| Lymphoid leukaemia | C91 | 3 | 232 | 5.31<br>(1.70 to 16.62) | 4.14[E-03] | 4 | 508 | 3.86<br>(1.44 to 10.31) | 7.20[E-03] | 4.42<br>(2.11 to 9.29) | 8.67[E-05] |
| Myeloid leukaemia | C92 | 0 | 126 |  |  | 0 | 361 |  |  |  |  |
| Carcinoma in situ of breast | D05 | 7 | 948 | 2.82<br>(1.33 to 5.96) | 6.70[E-03] | 9 | 1102 | 3.57<br>(1.85 to 6.88) | 1.44[E-04] | 3.22<br>(1.96 to 5.28) | 4.05[E-06] |
| Any cancer |  | 96 | 18742 | 2.14<br>(1.73 to 2.65) | 2.54[E-12] | 185 | 37617 | 2.34<br>(2.03 to 2.71) | < 2[E-16] | 2.28<br>(2.03 to 2.56) | 2.98[E-44] |
| Any cancer in females |  | 61 | 11774 | 2.10<br>(1.60 to 2.75) | 6.63[E-08] | 92 | 17261 | 2.32<br>(1.89 to 2.85) | 8.03 [E-16] | 2.23<br>(1.88 to 2.64) | 1.86[E-20] |
| Any cancer in females minus breast |  | 26 | 5534 | 1.82<br>(1.23 to 2.71) | 2.92[E-03] | 59 | 11498 | 2.30<br>(1.78 to 2.97) | 1.72[E-10] | 2.14<br>(1.73 to 2.65) | 2.79[E-12] |
| Any cancer in males |  | 35 | 6968 | 2.22<br>(1.57 to 3.14) | 7.47[E-06] | 93 | 19996 | 2.37<br>(1.93 to 2.90) | 2.00[E-16] | 2.33<br>(1.96 to 2.77) | 3.81[E-22] |
| Any cancer in males minus prostate |  | 25 | 4416 | 2.47<br>(1.64 to 3.71) | 1.32[E-05] | 52 | 11872 | 2.23<br>(1.70 to 2.93) | 7.30[E-09] | 2.30<br>(1.83 to 2.88) | 9.90[E-13] |

Pooled analyses also showed increased risk for any cancer in females (2·23 [1·88 to 2·64, p=1·86 x 10^−20^]) and males (2·33 [1·96 to 2·77, p=3·81 x 10^−22^]). These relative risks remained elevated (though slightly reduced in females), once breast and prostate cancer were excluded (2·14 [1·73 to 2·65, p=2·79 x 10^−12^] and (2·30 [1·83 to 2·88, p=9·90 x 10^−13^], respectively).

The OR and HR estimates from the retrospective and prospective analyses were similar for each of the associated cancers, and the differences were not statistically significant for any cancer type. The estimates for pancreatic cancer were higher in the retrospective analysis (20·64 [4·97 to 85·76, p=3·10 x 10^−5^]) than in the prospective analysis (6·15 [3·39 to 11·14, p=2·18 x 10^−9^]); however, carrier counts were low, and confidence intervals correspondingly wide.

There was some evidence of a decline in the HR with age for breast cancer (RR per decade 0·95 [0·92 to 0·99, p=0·012]) and all cancers in females (0·97 [0·95 to 0·99, p=0·013]), with the highest HR for breast cancer under age 50 years (3·92 [2·16 to 7·10, p=6·64 x 10^−6^]). There was no evidence for a trend in the HR by age for prostate or pancreatic cancer (Supplemental Table 1, Supplemental Table 5a).

### rMSVs in ATM

Pooled analyses (Table 2) showed that rMSV carriers had higher risks of cancer of the stomach (1·54 [95% CI: 1·07 to 2·23, p=0·022]); pancreas (1·60 [1·20 to 2·13, p=1·27 x 10^−3^]); prostate (1·15 [1·04 to 1·27, p=7·21 x 10^−3^]); and Hodgkin’s disease (HD) (1·72 [1·02 to 2·90, p=0·043]). There was some increased risk for any cancer males (1·13 [1·06 to 1·21, p=2·60 x 10^−4^]); removing prostate cancer made little difference to the estimate.

**Table 2:** Odds ratios, hazard ratios, and pooled estimates for the association of rMSVs in ATM with cancer by type.

| Cancer | ICD-10 | Carrier count | Non-carriers | OR (95% CI) | P-value | Carrier count | Non-carriers | HR (95% CI) | P-value | Pooled (95% CI) | P-value |
| --- | --- | --- | --- | --- | --- | --- | --- | --- | --- | --- | --- |
| Oesophagus | C15 | 2 | 112 | 0.50<br>(0.12 to 2.03) | 0.333 | 36 | 720 | 1.43<br>(1.03 to 2.00) | 0.035 | 1.35<br>(0.98 to 1.87) | 0.067 |
| Stomach | C16 | 6 | 100 | 1.67<br>(0.73 to 3.82) | 0.220 | 24 | 458 | 1.51<br>(1.00 to 2.27) | 0.050 | 1.54<br>(1.07 to 2.23) | 0.022 |
| Colon | C18 | 41 | 1014 | 1.13<br>(0.83 to 1.54) | 0.449 | 85 | 2677 | 0.90<br>(0.73 to 1.12) | 0.359 | 0.97<br>(0.81 to 1.16) | 0.746 |
| Rectum | C20 | 27 | 572 | 1.32<br>(0.90 to 1.94) | 0.162 | 36 | 1129 | 0.91<br>(0.65 to 1.26) | 0.559 | 1.06<br>(0.82 to 1.37) | 0.647 |
| Liver and intrahepatic bile ducts | C22 | 2 | 24 | 2.35<br>(0.56 to 9.97) | 0.245 | 19 | 427 | 1.27<br>(0.80 to 2.01) | 0.313 | 1.35<br>(0.86 to 2.11) | 0.190 |
| Pancreas | C25 | 3 | 38 | 2.17<br>(0.67 to 7.03) | 0.197 | 46 | 838 | 1.56<br>(1.16 to 2.10) | 3.19[E-03] | 1.60<br>(1.20 to 2.13) | 1.27[E-03] |
| Bronchus and lung | C34 | 9 | 233 | 1.08<br>(0.56 to 2.11) | 0.815 | 108 | 2622 | 1.18<br>(0.97 to 1.43) | 0.092 | 1.18<br>(0.98 to 1.42) | 0.090 |
| Melanoma of skin | C43 | 54 | 1445 | 1.04<br>(0.79 to 1.36) | 0.803 | 82 | 2095 | 1.11<br>(0.89 to 1.38) | 0.355 | 1.08<br>(0.91 to 1.28) | 0.373 |
| Breast | C50 | 229 | 6073 | 1.05<br>(0.91 to 1.20) | 0.517 | 236 | 5966 | 1.11<br>(0.97 to 1.26) | 0.128 | 1.07<br>(0.97 to 1.18) | 0.157 |
| Cervix uteri | C53 | 12 | 316 | 1.06<br>(0.59 to 1.89) | 0.847 | 2 | 84 | 0.68<br>(0.17 to 2.75) | 0.584 | 1.00<br>(0.59 to 1.69) | 0.992 |
| Corpus uteri | C54 | 24 | 654 | 1.01<br>(0.67 to 1.52) | 0.958 | 36 | 1016 | 0.99<br>(0.71 to 1.39) | 0.973 | 1.00<br>(0.77 to 1.29) | 0.987 |
| Ovary | C56 | 8 | 486 | 0.46<br>(0.23 to 0.92) | 0.028 | 27 | 659 | 1.15<br>(0.79 to 1.69) | 0.469 | 0.92<br>(0.66 to 1.30) | 0.650 |
| Prostate | C61 | 118 | 2444 | 1.35<br>(1.12 to 1.63) | 1.70[E-03] | 298 | 7867 | 1.08<br>(0.96 to 1.21) | 0.190 | 1.15<br>(1.04 to 1.27) | 7.21[E-03] |
| Testis | C62 | 10 | 396 | 0.70<br>(0.37 to 1.31) | 0.265 | 4 | 69 | 1.62<br>(0.59 to 4.43) | 0.351 | 0.89<br>(0.52 to 1.51) | 0.651 |
| Kidney, except renal pelvis | C64 | 15 | 373 | 1.12<br>(0.67 to 1.87) | 0.673 | 38 | 965 | 1.11<br>(0.81 to 1.54) | 0.513 | 1.12<br>(0.85 to 1.48) | 0.440 |
| Bladder | C67 | 18 | 414 | 1.21<br>(0.75 to 1.93) | 0.439 | 33 | 754 | 1.25<br>(0.88 to 1.77) | 0.214 | 1.23<br>(0.93 to 1.64) | 0.146 |
| Brain | C71 | 1 | 82 | 0.34<br>(0.05 to 2.43) | 0.282 | 17 | 540 | 0.89<br>(0.55 to 1.43) | 0.619 | 0.84<br>(0.52 to 1.35) | 0.470 |
| Thyroid | C73 | 10 | 260 | 1.08<br>(0.57 to 2.03) | 0.819 | 9 | 262 | 0.97<br>(0.50 to 1.88) | 0.921 | 1.02<br>(0.65 to 1.62) | 0.921 |
| Hodgkin's disease | C81 | 12 | 171 | 1.94<br>(1.08 to 3.49) | 0.027 | 3 | 79 | 1.08<br>(0.34 to 3.42) | 0.896 | 1.72<br>(1.02 to 2.90) | 0.043 |
| Diffuse non-Hodgkin's lymphoma | C83 | 12 | 286 | 1.16<br>(0.65 to 2.08) | 0.606 | 29 | 767 | 1.07<br>(0.74 to 1.55) | 0.712 | 1.10<br>(0.80 to 1.50) | 0.563 |
| Multiple myeloma and malignant plasma cell neoplasms | C90 | 3 | 142 | 0.58<br>(0.19 to 1.82) | 0.351 | 23 | 591 | 1.11<br>(0.73 to 1.68) | 0.632 | 1.03<br>(0.70 to 1.51) | 0.896 |
| Lymphoid leukaemia | C91 | 7 | 228 | 0.85<br>(0.40 to 1.81) | 0.679 | 22 | 490 | 1.28<br>(0.83 to 1.96) | 0.263 | 1.15<br>(0.79 to 1.67) | 0.464 |
| Myeloid leukaemia | C92 | 2 | 124 | 0.44<br>(0.11 to 1.79) | 0.253 | 15 | 346 | 1.23<br>(0.73 to 2.06) | 0.433 | 1.09<br>(0.68 to 1.77) | 0.714 |
| Carcinoma in situ of the breast | D05 | 37 | 918 | 1.12<br>(0.80 to 1.55) | 0.518 | 45 | 1066 | 1.18<br>(0.87 to 1.59) | 0.285 | 1.15<br>(0.92 to 1.43) | 0.220 |
| Any cancer |  | 701 | 18137 | 1.08<br>(1.00 to 1.16) | 0.063 | 1409 | 36393 | 1.10<br>(1.04 to 1.16) | 6.41[E-04] | 1.09<br>(1.04 to 1.14) | 5.61[E-04] |
| Any cancer in females |  | 417 | 11418 | 1.01<br>(0.91 to 1.12) | 0.823 | 664 | 17049 | 1.09<br>(1.01 to 1.18) | 0.024 | 1.06<br>(1.00 to 1.13) | 0.060 |
| Any cancer in females minus breast |  | 190 | 5370 | 0.98<br>(0.85 to 1.13) | 0.772 | 430 | 11127 | 1.09<br>(0.99 to 1.20) | 0.088 | 1.05<br>(0.97 to 1.15) | 0.221 |
| Any cancer in males |  | 284 | 6719 | 1.19<br>(1.05 to 1.34) | 6.15[E-03] | 745 | 19344 | 1.10<br>(1.02 to 1.19) | 9.49[E-03] | 1.13<br>(1.06 to 1.21) | 2.60[E-04] |
| Any cancer in males minus prostate |  | 166 | 4275 | 1.08<br>(0.93 to 1.27) | 0.314 | 447 | 11477 | 1.12<br>(1.02 to 1.23) | 0.023 | 1.11<br>(1.02 to 1.20) | 0.017 |

### rMSVs subset by functional protein domain and CADD score

Pooled analyses (Supplemental Table 7a-f) showed that RRs were highest for breast cancer (1·52 [95% CI: 1·14 to 2·04, p=4·85 x 10^−3^]) and prostate cancer (1·67 [1·23 to 2·29, p=1·20 x 10^−3^]) where variants lay inside the FAT or PIK domain, and had a CADD score within quintile 5 (>3·74) (compared with RRs for other variants: p-diff<0·0001 for both breast and prostate cancer). Similarly, the RRs for all cancers combined for variants in this category were higher than for all other variants combined, both in females (1·40 [95% CI: 1·15 to 1·71, p=8·74 x 10-4]; p-diff<0·0001); and males (1·34 [95% CI: 1·09 to 1·65, p=6·57 x 10^−3^]; p-diff<0·0001]).

For pancreatic cancer, the RR was highest where variants lay outside the FAT and PIK domains (1·56 [1·11 to 2·20, p=0·011]), but not significantly different to variants outside these domains (p-diff=0·65).

### PTVs in CHEK2

In the pooled analyses (Table 3), PTVs were associated with an increased risk of nine cancer types: oesophagus (RR: 2·13 [95% CI: 1·10 to 4·10, p=0·025]); melanoma 1·45 [1·00 to 2·09, p=0·049]); breast (2·44 [2·08 to 2·86, p=3·61 x 10^−28^]); ovary (2·33 [1·37 to 3·97, p=1·73 x 10^−3^]); prostate (1·92 [1·59 to 2·32, p=1·18 x 10^−11^]); kidney (1·83 [1·06 to 3·18, p=0·032]); HD (4·26 [1·89 to 9·64, p=4·93 x 10^−4^]); DNHL (1·97 [1·09 to 3·57, p=0·025]); and myeloid leukaemia (ML) (2·81 [1·33 to 5·94, p=6·75 x 10^−3^]). PTVs were also associated with CIS (2·05 [1·35 to 3·13, p=8·48 x 10^−4^]). The OR and HR estimates from the retrospective and prospective study were broadly similar for all the associated cancers, and did not differ significantly for any cancer type.

**Table 3:** Odds ratios, hazard ratios, and pooled estimates for the association of PTVs in CHEK2 with cancer by type.

| Cancer | ICD-10 | Carrier count | Non-carriers | OR (95% CI) | P-value | Carrier count | Non-carriers | HR (95% CI) | P-value | Pooled estimate (95% CI) | P-value |
| --- | --- | --- | --- | --- | --- | --- | --- | --- | --- | --- | --- |
| Oesophagus | C15 | 2 | 112 | 3.17<br>(0.78 to 12.85) | 0.107 | 7 | 749 | 1.89<br>(0.90 to 3.99) | 0.093 | 2.13<br>(1.10 to 4.10) | 0.025 |
| Stomach | C16 | 0 | 106 |  |  | 3 | 479 | 1.28<br>(0.41 to 3.98) | 0.671 | 1.01<br>(0.33 to 3.13) |  |
| Colon | C18 | 9 | 1046 | 1.54<br>(0.80 to 2.97) | 0.201 | 9 | 2753 | 0.65<br>(0.34 to 1.26) | 0.203 | 0.99<br>(0.62 to 1.57) | 0.957 |
| Rectum | C20 | 2 | 597 | 0.59<br>(0.15 to 2.35) | 0.451 | 5 | 1160 | 0.86<br>(0.36 to 2.06) | 0.731 | 0.77<br>(0.37 to 1.62) | 0.491 |
| Liver and intrahepatic bile ducts | C22 | 0 | 26 |  |  | 2 | 444 | 0.89<br>(0.22 to 3.59) | 0.874 | 0.84<br>(0.21 to 3.35) |  |
| Pancreas | C25 | 0 | 41 |  |  | 6 | 878 | 1.36<br>(0.61 to 3.04) | 0.452 | 1.29<br>(0.58 to 2.87) |  |
| Bronchus and lung | C34 | 0 | 242 |  |  | 14 | 2716 | 1.08<br>(0.64 to 1.83) | 0.772 | 0.98<br>(0.58 to 1.65) |  |
| Melanoma of skin | C43 | 11 | 1488 | 1.29<br>(0.71 to 2.35) | 0.396 | 17 | 2160 | 1.55<br>(0.96 to 2.49) | 0.074 | 1.45<br>(1.00 to 2.09) | 0.049 |
| Breast | C50 | 87 | 6215 | 2.65<br>(2.12 to 3.30) | <2[E-16] | 67 | 6135 | 2.23<br>(1.76 to 2.84) | 6.29[E-11] | 2.44<br>(2.08 to 2.86) | 3.61[E-28] |
| Cervix Uteri | C53 | 0 | 328 |  |  | 0 | 86 |  |  |  |  |
| Corpus uteri | C54 | 2 | 676 | 0.52<br>(0.13 to 2.08) | 0.354 | 7 | 1045 | 1.39<br>(0.66 to 2.93) | 0.383 | 1.12<br>(0.58 to 2.15) | 0.744 |
| Ovary | C56 | 6 | 488 | 2.18<br>(0.97 to 4.89) | 0.059 | 8 | 678 | 2.47<br>(1.23 to 4.95) | 0.011 | 2.33<br>(1.37 to 3.97) | 1.73[E-03] |
| Prostate | C61 | 27 | 2535 | 1.83<br>(1.25 to 2.69) | 2.09 [E-03] | 84 | 8081 | 1.96<br>(1.58 to 2.43) | 8.54[E-10] | 1.92<br>(1.59 to 2.32) | 1.18 [E-11] |
| Testis | C62 | 5 | 401 | 2.17<br>(0.89 to 5.25) | 0.087 | 0 | 73 |  |  | 1.82<br>(0.76 to 4.38) |  |
| Kidney, except renal pelvis | C64 | 5 | 383 | 2.29<br>(0.95 to 5.54) | 0.067 | 8 | 995 | 1.58<br>(0.79 to 3.17) | 0.197 | 1.83<br>(1.06 to 3.18) | 0.032 |
| Bladder | C67 | 3 | 429 | 1.17<br>(0.38 to 3.66) | 0.784 | 5 | 782 | 1.25<br>(0.52 to 3.00) | 0.624 | 1.22<br>(0.61 to 2.45) | 0.579 |
| Brain | C71 | 2 | 81 | 4.28<br>(1.05 to 17.45) | 0.043 | 0 | 557 |  |  | 0.61<br>(0.15 to 2.43) |  |
| Thyroid | C73 | 3 | 267 | 2.07<br>(0.66 to 6.47) | 0.211 | 1 | 270 | 0.76<br>(0.11 to 5.41) | 0.783 | 1.61<br>(0.60 to 4.30) | 0.343 |
| Hodgkin's disease | C81 | 4 | 179 | 3.88<br>(1.44 to 10.48) | 7.39[E-03] | 2 | 80 | 5.11<br>(1.25 to 20.82) | 0.023 | 4.26<br>(1.89 to 9.64) | 4.93[E-04] |
| Diffuse non-Hodgkin's lymphoma | C83 | 4 | 294 | 2.38<br>(0.89 to 6.39) | 0.086 | 7 | 789 | 1.77<br>(0.84 to 3.73) | 0.132 | 1.97<br>(1.09 to 3.57) | 0.025 |
| Multiple myeloma and malignant cell neoplasms | C90 | 1 | 144 | 1.21<br>(0.17 to 8.62) | 0.853 | 3 | 611 | 0.97<br>(0.31 to 3.02) | 0.961 | 1.03<br>(0.38 to 2.74) | 0.960 |
| Lymphoid leukaemia | C91 | 0 | 235 |  |  | 7 | 505 | 2.73<br>(1.29 to 5.75) | 8.49[E-03] | 1.78<br>(0.85 to 3.74) |  |
| Myeloid leukaemia | C92 | 3 | 123 | 4.07<br>(1.29 to 12.82) | 0.017 | 4 | 357 | 2.17<br>(0.81 to 5.81) | 0.125 | 2.81<br>(1.33 to 5.94) | 6.75[E-03] |
| Carcinoma in situ of breast | D05 | 11 | 944 | 2.10<br>(1.16 to 3.82) | 0.015 | 11 | 1100 | 2.01<br>(1.11 to 3.63) | 0.022 | 2.05<br>(1.35 to 3.13) | 8.48[E-04] |
| Any cancer |  | 192 | 18646 | 1.91<br>(1.64 to 2.21) | <2[E-16] | 297 | 37505 | 1.57<br>(1.40 to 1.76) | 8.58[E-15] | 1.68<br>(1.53 to 1.85) | 3.44[E-27] |
| Any cancer in females |  | 122 | 11713 | 1.99<br>(1.64 to 2.40) | 1.55[E-12] | 141 | 17572 | 1.67<br>(1.41 to 1.97) | 1.59[E-09] | 1.81<br>(1.58 to 2.06) | 1.07[E-18] |
| Any cancer in females minus breast |  | 36 | 5524 | 1.18<br>(0.84 to 1.64) | 0.345 | 74 | 11483 | 1.35<br>(1.07 to 1.70) | 0.010 | 1.29<br>(1.06 to 1.56) | 9.73[E-03] |
| Any cancer in males |  | 70 | 6933 | 1.78<br>(1.40 to 2.28) | 3.57[E-06] | 156 | 19933 | 1.50<br>(1.28 to 1.75) | 5.05[E-07] | 1.57<br>(1.37 to 1.79) | 4.21[E-11] |
| Any cancer in males minus prostate |  | 43 | 4398 | 1.71<br>(1.26 to 2.33) | 6.17[E-04] | 72 | 11852 | 1.18<br>(0.93 to 1.48) | 0.174 | 1.35<br>(1.12 to 1.62) | 1.99[E-03] |

The risk of all cancers combined was elevated in both females (1·81 [1·58 to 2·06, p=1·07 x 10^−18^]), and males (1·57 [1·37 to 1·79, p=4·21 x 10^−11^]). After excluding breast cancer in females and prostate cancer in males, the RRs were reduced but remained elevated (1·29 [1·06 to 1·56, p=9·73 x 10^−3^], and 1·35 [95% CI: 1·12 to 1·62, p=1·99 x 10^−3^], respectively).

We examined the variation in the HR by age for breast, prostate, pancreatic, and any cancer. There was evidence of association in all age-groups for breast and prostate cancer, and no evidence of differences in the HR by age for any cancer (Supplemental Table 3, Supplemental Table 6a).

### rMSVs in CHEK2

In the pooled analyses (Table 4), rMSVs were associated with a higher risk of five cancer types: melanoma (1·32 [95% CI: 1·01 to 1·71, p=0·041]); breast cancer (1·50 [1·31 to 1·72, p=1·02 x 10^−8^]); prostate cancer (1·41 [1·21 to 1·64, p=1·43 x 10^−5^]); multiple myeloma (1·86 [1·13 to 3·05, p=0·014]); and LL (2·23 [1·42 to 3·52, p=5·22 x 10^−4^]). rMSVs also were associated with CIS (1·55 [1·13 to 2·14, p=6·82 x 10^−3^]).

**Table 4:** Odds ratios, hazard ratios, and pooled estimates for the association of rMSVs in CHEK2 with cancer by type.

| Cancer | ICD-10 | Carrier count | Non-carrier | OR (95% CI) | P-value | Carrier count | Non-carriers | HR (95% CI) | P-value | Pooled estimate (95% CI) | P-value |
| --- | --- | --- | --- | --- | --- | --- | --- | --- | --- | --- | --- |
| Oesophagus | C15 | 0 | 114 |  |  | 6 | 750 | 0.70<br>(0.31 to 1.56) | 0.385 | 0.61<br>(0.27 to 1.35) |  |
| Stomach | C16 | 0 | 106 |  |  | 3 | 479 | 0.55<br>(0.18 to 1.72) | 0.305 | 0.45<br>(0.15 to 1.40) |  |
| Colon | C18 | 15 | 1040 | 1.19<br>(0.71 to 1.98) | 0.515 | 35 | 2727 | 1.11<br>(0.80 to 1.55) | 0.529 | 1.14<br>(0.86 to 1.50) | 0.369 |
| Rectum | C20 | 10 | 589 | 1.40<br>(0.75 to 2.61) | 0.297 | 11 | 1154 | 0.82<br>(0.45 to 1.49) | 0.513 | 1.06<br>(0.58 to 1.92) | 0.855 |
| Liver and intrahepatic bile ducts | C22 | 0 | 26 |  |  | 5 | 441 | 1.01<br>(0.42 to 2.43) | 0.986 | 0.95<br>(0.40 to 2.28) |  |
| Pancreas | C25 | 1 | 40 | 2.03<br>(0.28 to 14.81) | 0.484 | 10 | 874 | 0.99<br>(0.53 to 1.85) | 0.979 | 1.06<br>(0.58 to 1.92) | 0.855 |
| Bronchus and lung | C34 | 3 | 239 | 1.05<br>(0.34 to 3.27) | 0.939 | 26 | 2704 | 0.84<br>(0.57 to 1.24) | 0.388 | 0.86<br>(0.60 to 1.25) | 0.438 |
| Melanoma of skin | C43 | 27 | 1472 | 1.48<br>(1.01 to 2.17) | 0.044 | 30 | 2147 | 1.19<br>(0.83 to 1.71) | 0.343 | 1.32<br>(1.01 to 1.71) | 0.041 |
| Breast | C50 | 112 | 6190 | 1.46<br>(1.20 to 1.77) | 1.16[E-04] | 113 | 6089 | 1.54<br>(1.28 to 1.85) | 6.30[E-06] | 1.50<br>(1.31 to 1.72) | 1.02[E-08] |
| Cervix uteri | C53 | 5 | 323 | 1.24<br>(0.51 to 3.01) | 0.629 | 0 | 86 |  |  | 1.01<br>(0.42 to 2.44) |  |
| Corpus uteri | C54 | 8 | 670 | 0.93<br>(0.46 to 1.88) | 0.847 | 14 | 1038 | 1.12<br>(0.66 to 1.89) | 0.682 | 1.05<br>(0.69 to 1.60) | 0.834 |
| Ovary | C56 | 8 | 486 | 1.31<br>(0.65 to 2.63) | 0.454 | 8 | 678 | 0.98<br>(0.49 to 1.97) | 0.959 | 1.13<br>(0.69 to 1.87) | 0.623 |
| Prostate | C61 | 41 | 2521 | 1.37<br>(1.00 to 1.87) | 0.051 | 130 | 8035 | 1.42<br>(1.19 to 1.69) | 7.28[E-05] | 1.41<br>(1.21 to 1.64) | 1.43 [E-05] |
| Testis | C62 | 7 | 399 | 1.48<br>(0.70 to 3.14) | 0.302 | 1 | 72 | 1.16<br>(0.16 to 8.36) | 0.882 | 1.43<br>(0.71 to 2.88) | 0.311 |
| Kidney, except renal pelvis | C64 | 3 | 385 | 0.64<br>(0.21 to 1.99) | 0.438 | 14 | 989 | 1.21<br>(0.71 to 2.05) | 0.481 | 1.08<br>(0.67 to 1.74) | 0.756 |
| Bladder | C67 | 10 | 422 | 1.96<br>(1.05 to 3.68) | 0.036 | 10 | 777 | 1.12<br>(0.60 to 2.09) | 0.718 | 1.48<br>(0.95 to 2.31) | 0.081 |
| Brain | C71 | 0 | 83 |  |  | 4 | 553 | 0.61<br>(0.23 to 1.64) | 0.329 | 0.53<br>(0.20 to 1.42) |  |
| Thyroid | C73 | 5 | 265 | 1.52<br>(0.63 to 3.70) | 0.351 | 3 | 268 | 0.93<br>(0.30 to 2.91) | 0.906 | 1.27<br>(0.63 to 2.54) | 0.507 |
| Hodgkin's disease | C81 | 3 | 180 | 1.35<br>(0.43 to 4.23) | 0.605 | 1 | 81 | 1.06<br>(0.15 to 7.62) | 0.953 | 1.27<br>(0.48 to 3.41) | 0.633 |
| Diffuse non-Hodgkin's lymphoma | C83 | 1 | 297 | 0.28<br>(0.04 to 1.96) | 0.199 | 9 | 787 | 0.99<br>(0.51 to 1.91) | 0.974 | 0.87<br>(0.46 to 1.63) | 0.658 |
| Multiple myeloma and malignant plasma cell neoplasms | C90 | 2 | 143 | 1.14<br>(0.28 to 4.59) | 0.858 | 14 | 600 | 1.99<br>(1.17 to 3.39) | 0.011 | 1.86<br>(1.13 to 3.05) | 0.014 |
| Lymphoid leukaemia | C91 | 5 | 230 | 1.80<br>(0.74 to 4.37) | 0.195 | 14 | 498 | 2.41<br>(1.41 to 4.09) | 1.21[E-03] | 2.23<br>(1.42 to 3.52) | 5.22[E-04] |
| Myeloid leukaemia | C92 | 0 | 126 |  |  | 8 | 353 | 1.95<br>(0.97 to 3.93) | 0.062 | 1.40<br>(0.70 to 2.81) |  |
| Carcinoma in situ of breast | D05 | 20 | 935 | 1.69<br>(1.09 to 2.64) | 0.020 | 19 | 1092 | 1.42<br>(0.90 to 2.24) | 0.127 | 1.55<br>(1.13 to 2.14) | 6.82[E-03] |
| Any cancer |  | 304 | 18534 | 1.35<br>(1.20 to 1.52) | 5.01[E-07] | 535 | 37267 | 1.23<br>(1.13 to 1.34) | 2.69[E-06] | 1.26<br>(1.18 to 1.35) | 4.10[E-12] |
| Any cancer in females |  | 200 | 11635 | 1.40<br>(1.21 to 1.62) | 7.75[E-06] | 253 | 17460 | 1.20<br>(1.06 to 1.36) | 3.83[E-03] | 1.26<br>(1.15 to 1.39) | 1.09[E-06] |
| Any cancer in females minus breast |  | 88 | 5472 | 1.28<br>(1.04 to 1.59) | 0.023 | 141 | 11416 | 1.03<br>(0.87 to 1.21) | 0.772 | 1.13<br>(0.98 to 1.29) | 0.090 |
| Any cancer in males |  | 104 | 6899 | 1.28<br>(1.05 to 1.56) | 0.016 | 282 | 19807 | 1.26<br>(1.12 to 1.42) | 1.20[E-04] | 1.27<br>(1.14 to 1.40) | 4.80[E-06] |
| Any cancer in males minus prostate |  | 63 | 4378 | 1.22<br>(0.94 to 1.56) | 0.132 | 152 | 11772 | 1.15<br>(0.98 to 1.35) | 0.091 | 1.17<br>(1.02 to 1.34) | 0.022 |

Combined estimates also showed increased risk of any cancer in females (1·26 [1·15 to 1·39, p=1·09 x 10^−6^]), but the risk was no longer significant after excluding breast cancer (1·13 [0·98 to 1·29, p=0·090]). Risk of any cancer was also elevated for males (1·27 [1·14 to 1·40, p=4·80 x 10^−6^]) and after exclusion of prostate cancer (1·17 [1·02 to 1·34, p=0·022]).

Statistically significant risk estimates from the retrospective and prospective analyses were broadly similar, and the differences were not statistically significant for any cancer type.

### rMSVs Helix score

rMSVs were categorised into high (>0·5) or low (<=0·5) Helix scores (Supplemental Tables 8a-f). In the pooled analyses, the RR for any cancer in females was higher in the Helix-high group (1·43 [95% CI: 1·23 to 1·65, p=1·60 x 10^−6^]) than in the Helix-low group (1·20 [1·06 to 1·36, p=3·72 x 10^−3^]); p-diff<0·0001]). A higher RR for breast cancer was also found in the Helix-high group (1·62 [95% CI: 1·31 to 2·01, p=9·53 x 10^−6^] versus (1·42 [95% CI: 1·20 to 1·69 p=6·37 x 10^−5^]; p-diff=0·031) in the Helix-low group. A similar pattern was observed for any cancer in males (Helix-high: 1·35 [1·15 to 1·60, p=3·61 x 10^−4^]), and (Helix-low: 1·22 [1·06 to 1·39, p=4·31 x 10^−3^]); p-diff=4·34 x 10^−4^]), but not for prostate cancer.

### Absolute risks

Supplemental Figure 1 shows the cumulative risks of female breast cancer by gene and variant type. By age 80, the cumulative risk of breast cancer was 30% (95% CI: 20 to 38%) for *ATM* PTV carriers and 25% (20 to 29%) for *CHEK2* PTV carriers, compared with 12% in the general population. The corresponding average cumulative risks for rMSV carriers were 18% (15 to 20%) for *CHEK2* and 13% (11 to 15%) for *ATM* .

For prostate cancer (Supplemental Figure 2), the cumulative risks by age 80 were 31% (23 to 38%) for *ATM* PTV carriers and 25 (20 to 29%) for *CHEK2* PTV carriers. The corresponding average cumulate risk for rMSV carriers was 19% (0·16 to 0·21%) for *CHEK2* and 14% (13 to 0·16%) for *ATM* .

The risks of pancreatic cancer for *ATM* mutation carriers by sex and variant type are shown in Figure 2. By age 80, the estimated cumulative risk of pancreatic cancer in *ATM* PTV carriers was 8% (2 to 15%) in males and 6% (1 to 10%) in females. The cumulative risks for rMSV carriers were 2% (1 to 3%) in males, and 2% (1 to 2%) in females.

**Figure 2:**
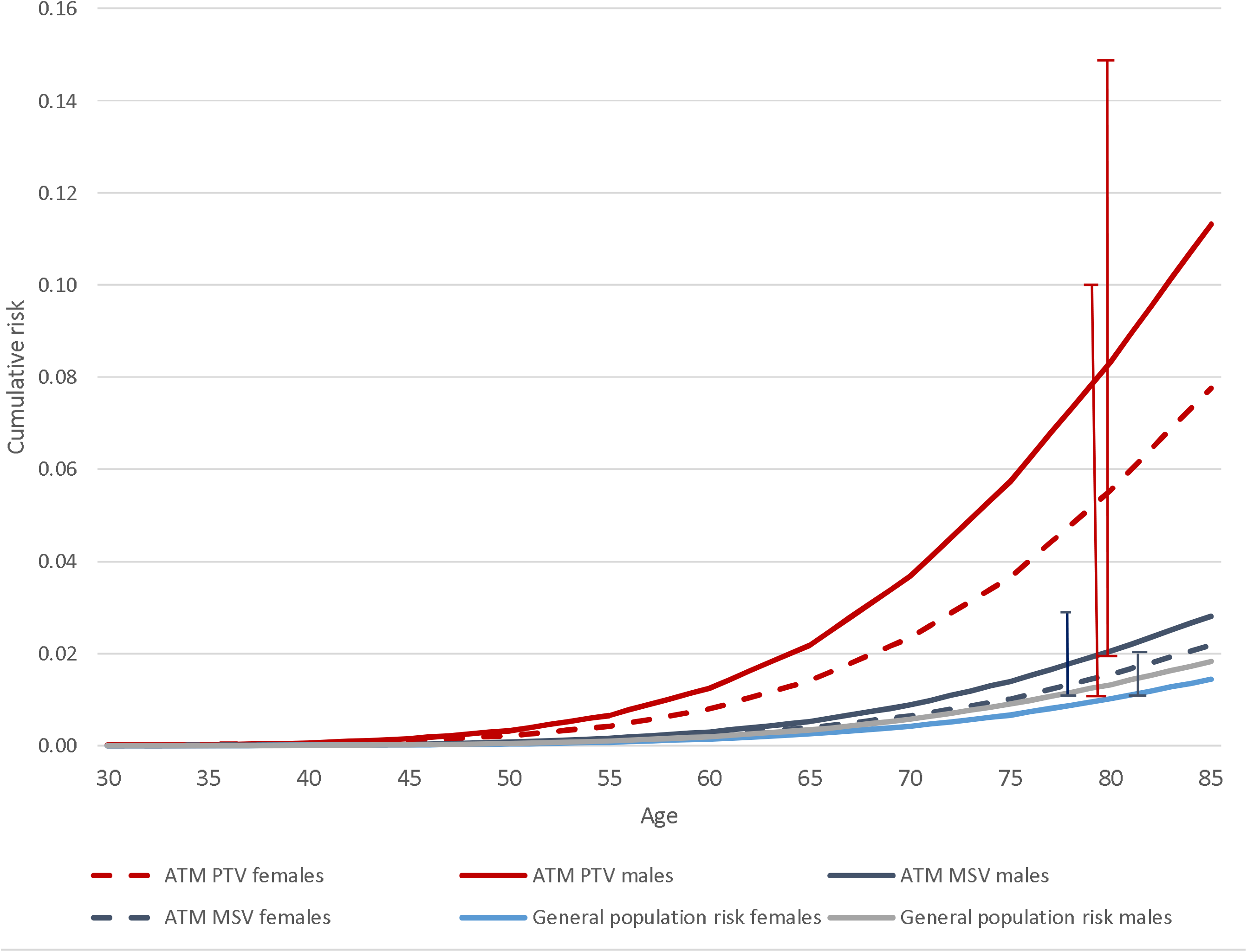
Cumulative absolute pancreatic cancer risks for *ATM* protein-truncating/missense variant carriers and the general population by age and sex

The cumulative risk of developing any cancer by the age of 80 for *ATM* PTV carriers was 59% (95% CI: 50 to 67%) for females and 69 (61 to 76%) for males. The corresponding cumulative risks for *CHEK2* PTV carriers was 49% (43 to 54%) for females and 52% (46 to 58%) for males (Supplemental Figure 3, Supplemental Figure 4, respectively).

## Discussion

The pooled analyses show clear evidence of association between carrying a PTV in either gene and a higher risk of multiple cancers. For *ATM*, the RR for breast cancer (2·27 in the combined analysis) was consistent with previous case-control and family-based analyses (6). The RR for CIS (3·22 [95% CI: 1·96-5·28]) was consistent with that for invasive breast cancer, and a similar RR was found for prostate cancer (2·35). The excess risk of other (non-breast and prostate) cancer was more marked than for *CHEK2* PTVs, with a pooled RR of approximately two-fold in both males and females. *ATM* PTVs were associated with significantly increased risk for 11 of 23 sites examined (9 at p<0·001), and (as for CHEK2), no sites had a significantly reduced risk. A previously reported association for pancreatic cancer was confirmed (30), with the RR for pancreatic cancer being the highest for any cancer type. There was some similarity with the spectrum of cancers associated with *CHEK2* PTVs (e.g., oesophageal cancer, DNHL, and melanoma being common), presumably reflecting their related roles in DNA damage response; notable differences are of the association with colon and lung cancer in *ATM* PTV carriers not seen in *CHEK2*.

For *ATM*, rMSVs, there was only a small excess cancer risk overall (RRs 1·06 in females and 1·13 in males). Previous research has shown that breast cancer risk appears to be largely restricted to a subset of evolutionary conserved missense variants in the FAT and PIK domains (9). When analyses were restricted to these variants, risks for breast cancer, prostate cancer, any cancer in females, and any cancer in males, were higher for variants which lay within the FAT or PIK domains, with a high CADD score, and largely restricted to these variants. For pancreatic cancer, there was no evidence of a difference in RR by *in-silico* risk category, but the confidence limits were wide.

For *CHEK2* PTVs, the estimated RR for breast cancer (approximately two-fold) is similar to that previously reported for recent large case-control studies (6, 8). The RR for prostate cancer (1·92) is somewhat lower than some previous estimates (19, 40), but here we provide much stronger evidence for the association with prostate cancer risk, and more precise estimates. The association with CIS, with a RR similar to that for invasive disease, is also consistent with that reported previously for c.1100delC (39).

There was also evidence for an excess risk of other cancers in aggregate in both males and females. While breast and prostate cancer explained more than 50% of this excess, these excess risks remained statistically significant after excluding these cancers. Statistically significant increased risks were observed for 7 of the other 21 sites considered: oesophagus, ovary, kidney, HD, DNHL, ML, and melanoma. It is notable that no common sites were associated with a reduced risk—this and the clear association overall cancer risk, after excluding breast and prostate cancer, suggests that many of these associations are genuine. While some other sites (e.g., kidney) have been previously suggested to be associated (19), none have been firmly established. We did not, however, find evidence to support the previous observation of an increased risk of colorectal cancer

Rare MSVs in *CHEK2* were also associated with increased risk of both breast and prostate cancer, with RRs of approximately 1·4, comparable to previous estimates for breast cancer (6, 25), and to estimates for the association of the I157T variant and prostate cancer (10, 40). The RR for breast cancer was higher for variants classified as likely deleterious by Helix score. These results confirm, with similar RR estimates, previous analyses showing that Helix scores are predictive of risk, but that variants with low scores are also associated with risk. A similar pattern was seen for all cancers in both males and females, but not for prostate cancer. The latter result might, however, be due to chance, given the results for all cancers combined. A significant excess risk in rMSV carriers was observed for several other cancer types (melanoma, multiple myeloma, and LL), which were also associated with PTVs. Taken together, this is broadly consistent with the hypothesis that *CHEK2* MSVs confer increased, but on average lower, risks of a similar spectrum of cancers to PTVs, with risks being mutation dependent. High-throughput functional assays to classify the degree of *CHEK2* abrogation are becoming available and may be able to define variant-specific risks more precisely (41-43). Previous studies have shown that the CHEK2*I157T variant is associated with a reduced risk for lung cancer (44). There were too few carriers of this variant to investigate individually, but there was no evidence of a reduced risk associated with rMSVs in aggregate for this site.

### Strengths and limitations

A major strength of this study is that, as a very large cohort study, it was possible to assess the risks for many cancer types simultaneously. Moreover, as individuals were genotyped independently of phenotype, the analyses should free from biases due to differential genotyping of cases and controls. In contrast to family-based studies, the risk estimates should be broadly applicable to the population. A potential limitation is that UK Biobank, in common with similar cohorts, is biased towards healthy volunteers: total cancer incidence at age 70-74 is 12-18% lower than in the general population (45). However, while this affects the power to detect associations, it is unlikely that the RRs will be materially biased. Further, we generated absolute risk estimates by applying these RRs to national incidence rates. A significant limitation is that more than 90% of the cohort is of European ancestry, and we therefore restricted the analyses to this population. Other analyses indicate that the RRs conferred by cancer susceptibility genes tend to be similar across populations of different ancestries (6, 46); nevertheless, extrapolation to other ancestries may be less reliable (particularly if there are ancestry-specific variants associated with different risks).

### Implications

*ATM* and *CHEK2* are included on many cancer gene panels used in family cancer clinics, and the risk estimates from these analyses can inform genetic counselling for carriers. The estimated absolute risks for pancreatic cancer in *ATM* PTV carriers (11% in males and 8% in females by age 85) are notably higher than for other major pancreatic susceptibility genes including BRCA2, CDK2NA, and PALB2, though the confidence limits are wide. Current NICE guidelines for pancreatic cancer management do not include *ATM*, though it is included in other guidelines. Given the frequency of *ATM* variants, *ATM* PTVs would explain ∼2% of pancreatic cancer cases (with potentially a higher proportion due to MSVs). Prognosis for pancreatic cancer is poor, with only 7% of patients diagnosed with pancreatic cancer between 2013 and 2017 in England surviving 5 years (47). *ATM* variant carriers may provide a suitable population in which to evaluate new methods of early detection of this disease. Moreover, given the high excess lifetime risk for all cancers, carriers may a good target population for methods, such as those based on circulating tumour DNA, than can be utilised for multiple cancer types (48).

It is important to note that cancer risks are determined by a combination of multiple factors, including rare gene variants such as those in *ATM* and *CHEK2* studied here, but also lifestyle factors, and commoner genetic variants. Ideally, counselling should be based on synthesizing all available data, using risk models such as BOADICEA/CanRisk that have been developed for breast, ovarian, and prostate cancer. An ongoing challenge is to extend such models to consider a wider range of cancers.

Our analyses demonstrate that *ATM* and *CHEK2* PTVs are associated with increased risks of a wide range of cancers, with the overall cancer risk being higher for *ATM*. *CHEK2* rMSVs are also associated with smaller risks of a similar range of cancers; for *ATM* (but not CHEK2) the risks appear to be restricted to a small subset of rMSVs. The absolute magnitudes of these risks (with more than half of PTV carriers being affected by age 80) are significant and may influence guidelines for counselling and management.

## Data Availability

Requests for access to UK Biobank data should be made to the UK Biobank Access Management Team.

## Funding and Acknowledgments

This work has been funded by European Union’s Horizon 2020 Research and Innovation Program (BRIDGES grant 634935), Wellcome Trust (grant v203477/Z/16/Z), Cancer Research UK grant (PPRPGM-Nov20\100002), and the Medical Research Council (unit program: MC_UU_12015/2,MC_UU_00006/2). This work was also supported by core funding from the NIHR Cambridge Biomedical Research Centre (NIHR203312). The views expressed are those of the author(s) and not necessarily those of the NIHR or the Department of Health and Social Care.

NW was supported by the International Alliance for Cancer Early Detection, an alliance between Cancer Research UK (C14478/A29329), Canary Center at Stanford University, the University of Cambridge, OHSU Knight Cancer Institute, University College London, and the University of Manchester.

## Ethics approval

UK Biobank has approval from the North West Multi-centre Research Ethics Committee (MREC). Research Ethics Committee reference 11/NW/0382). This research has been conducted using the UK Biobank Resource under Application Number 28126.

## Authors’ contributions

DFE conceived the study, supervised the work, and directed the overall analyses. TKM performed the statistical analyses. TKM and DFE drafted the manuscript. JRBP, EJG, and NW developed the bioinformatics pipeline for variant calling and quality control. NW, NM, JD, and XY provided data management support. MN provide bioinformatics support. All authors reviewed and approved the paper.

## Data sharing

Requests for access to UK Biobank data should be made to the UK Biobank Access Management Team.

