## Supplemental Material for "Protein-truncating and rare missense variants in *ATM* and *CHEK2* and associations with cancer in UK Biobank whole-exome sequenced data"

*Variant filtering, classification, and functional prediction*

Quality Control (QC) metrics that have been previously documented were applied to VCF files (1, 2). Variants were annotated using the Ensembl Variant Effect Predictor (VEP), version 101, using the MANE Select transcript for each gene (3). Frameshift, splice acceptor, splice donor, stop gained, and start lost variants were grouped as PTVs. PTVs in the last exon were excluded, as were those in the last 50 base positions of the penultimate exon, to exclude variants likely to avoid Nonsense-Mediated mRNA Decay (4). In *ATM*, rMSVs were categorised by functional protein domain (5) and pathogenicity score predicted by the Combined Annotation Dependent Depletion algorithm (CADD; version 1.6) (6). In *CHEK2*, rMSVs were classified as high or low-risk using the deleteriousness score given by the Helix algorithm (version; 4.4.1) (7). These classifications were used as they provided the best discrimination for breast cancer risk in the analysis of the large BRIDGES dataset and hence the analysis provided a basis for replicating those findings (8).

Participants with unknown genetic sex, genetic sex that did not match reported sex, or sex chromosome aneuploidies, were excluded from the analyses. Participants of non-European ancestry, identified either by genetic ancestry (defined using principal components analysis) (9) or self-reported ancestry, were also excluded.

To avoid complications with inclusion of closely related individuals, one of each related pair of first-degree relatives (where the kinship coefficient >0·17) was excluded, as were participants estimated to have >10 relatives up to third-degree in the cohort.

*Combined Relative Risk Estimates*

Under the rare disease assumption, the OR and HR should both estimate the same underlying rate ratio or relative risk (RR). Thus, it is possible combine the estimates from the retrospective and prospective analyses in meta-analysis. The equivalence of these estimates also depends on additional assumptions that there is no association between the variants and survival, and no differences in the RR by age. However, even if this is not exactly true, the meta-analysis should still provide a more powerful test of association.

Thus, for each gene, fixed effects meta-analyses of the ORs from the retrospective study, and HRs from the prospective study, were conducted using the log of each ratio and its standard error. Where there were zero carriers in cases in either the retrospective or prospective studies, a combined estimate was derived as (OR+OP)/(ER+EP), where OR and OP are the observed number of carriers in cases in the retrospective and prospective analysis, and ER and EP are the corresponding predicted numbers of carriers, using the predict function in R applied to the logistic or Cox regression models but without carrier status. The corresponding standard error of the log(RR) estimate is then approximately 1/sqrt(OR+OP). This method should be reasonably accurate when the (combined carrier) frequency is low, as here, since then the observed counts then follow a Poisson distribution.
