## Supplemental Tables for "Protein-truncating and rare missense variants in *ATM* and *CHEK2* and associations with cancer in UK Biobank whole-exome sequenced data"

Supplemental Table 1: Cox regression analyses for the association of PTVs in ATM with cancer by type, using a continuous time-dependent coefficient

Supplemental Table 2: Cox regression analyses for the association of rMSVs in ATM with cancer by type using a continuous time-dependent coefficient

Supplemental Table 3: Cox regression analyses for the association of PTVs in CHEK2 with cancer by type, using a continuous time-dependent coefficient

Supplemental Table 4: Cox regression analyses for the association of rMSVs in CHEK2 with cancer by type, using a continuous time-dependent coefficient

Supplemental Table 5a: Cox regression analyses for the association of PTVs in ATM with cancer by type using a time-dependent coefficient

Supplemental Table 5b: Cox regression analyses for the association of rMSVs in ATM with cancer by type using a time-dependent coefficient

Supplemental Table 6a: Cox regression analyses for the association of PTVs in CHEK2 with cancer by type using a time-dependent coefficient

Supplemental Table 6b: Cox regression analyses for the association of rMSVs in CHEK2 with cancer by type using a time-dependent coefficient

Supplemental Table 7a: Odds ratios and hazard ratios for the association of rMSVs in ATM with pancreatic cancer, by domain and CADD score

Supplemental Table 7b: Odds ratios and hazard ratios for the association of rMSVs in ATM with breast cancer, by domain and CADD score

Supplemental Table 7c: Odds ratios and hazard ratios for the association of rMSVs in ATM with prostate cancer, by domain and CADD score

Supplemental Table 7d: Odds ratios and hazard ratios for the association of rMSVs in ATM with any cancer, by domain and CADD score

Supplemental Table 7e: Odds ratios and hazard ratios for the association of rMSVs in ATM with any cancer in females, by domain and CADD score

Supplemental Table 7f: Odds ratios and hazard ratios for the association of rMSVs in ATM with any cancer in males, by domain and CADD score

Supplemental Table 8a: Odds ratios and hazard ratios for the association of rMSVs in CHEK2 with breast cancer by Helix score

Supplemental Table 8b: Odds ratios and hazard ratios for the association of rMSVs in CHEK2 with prostate cancer by Helix score

Supplemental Table 8c: Odds ratios and hazard ratios for the association of rMSVs in CHEK2 with pancreatic cancer by Helix score

Supplemental Table 8d: Odds ratios and hazard ratios for the association of rMSVs in CHEK2 with any cancer in males and females by Helix score

Supplemental Table 8e: Odds ratios and hazard ratios for the association of rMSVs in CHEK2 with any cancer in females by Helix score

Supplemental Table 8f: Odds ratios and hazard ratios for the association of rMSVs in CHEK2 with any cancer in males by Helix score

***Supplemental Table 1: Cox regression analyses for the association of PTVs in ATM with cancer by type, using a continuous time-dependent coefficient***

|  | **HR (95% CI)** | ***P-*value** |
| --- | --- | --- |
| **Breast** |  |  |
| variant | 4·08 (2·49 to 6·68) | 2·25[E-08] |
| variant x age^1^ | 0·95 (0·92 to 0·99) | 0·012 |
| **Prostate** |  |  |
| variant | 2·25 (0·82 to 6·23) | 0·118 |
| variant x age^1^ | 1·01 (0·96 to 1·06) | 0·796 |
| **Pancreatic** |  |  |
| variant | 4·30 (0·72 to 25·83) | 0·110 |
| variant x age^1^ | 1·02 (0·93 to 1·11) | 0·672 |
| **Any cancer females** |  |  |
| variant | 3·49 (2·43 to 5·01) | 1·14[E-11] |
| variant x age^1^ | 0·97 (0·95 to 0·99) | 0·013 |
| **Any cancer males** |  |  |
| variant | 2·85 (1·64 to 4·95) | 2·11[E-04] |
| variant x age^1^ | 0·99 (0·96 to 1·02) | 0·487 |

^1^Fit using a model of the form coxph(Surv(time,event)~variant+tt(variant)+…,tt = function(x,t, ...) x*(t-50)). The main effect is thus the estimated HR associated with carrying a variant at age 50 years. The interaction term is the estimated relative change in the HR associated with carrying a variant, per year.

***Supplemental Table 2: Cox regression analyses for the*** ***association of rMSVs in ATM with cancer by type using a continuous time-dependent coefficient***

|  | **HR (95% CI)** | ***P-*value** |
| --- | --- | --- |
| **Breast** |  |  |
| variant | 1·17 (0·91 to 1·50) | 0·211 |
| variant x age^1^ | 1·00 (0·98 to 1·01) | 0·593 |
| **Prostate** |  |  |
| variant | 1·16 (0·80 to 1·68) | 0·443 |
| variant x age^1^ | 1·00 (0·98 to 1·02) | 0·705 |
| **Pancreatic** |  |  |
| variant | 1·40 (0·59 to 3·34) | 0·451 |
| variant x age^1^ | 1·01 (0·96 to 1·05) | 0·788 |
| **Any cancer females** |  |  |
| variant | 1·19 (1·01 to 1·41) | 0·036 |
| variant x age^1^ | 0·99 (0·99 to 1·00) | 0·249 |
| **Any cancer males** |  |  |
| variant | 1·06 (0·86 to 1·31) | 0·602 |
| variant x age^1^ | 1·00 (0·99 to 1·01) | 0·688 |

^1^Fit using a model of the form coxph(Surv(time,event)~variant+tt(variant)+…,tt = function(x,t, ...) x*(t-50)). The main effect is thus the estimated HR associated with carrying a variant at age 50 years. The interaction term is the estimated relative change in the HR associated with carrying a variant, per year.

***Supplemental Table 3: Cox regression analyses for the association of PTVs in CHEK2 with cancer by type, using a continuous time-dependent coefficient***

|  | **HR (95% CI)** | ***P-*value** |
| --- | --- | --- |
| **Breast** |  |  |
| variant | 2·41 (1·55 to 3·74) | 9·42[E-05] |
| variant x age^1^ | 1·00 (0·97 to 1·02) | 0·725 |
| **Prostate** |  |  |
| variant | 3·14 (1·66 to 5·95) | 4·41[E-04] |
| variant x age^1^ | 0·97 (0·94 to 1·01) | 0·135 |
| **Pancreatic** |  |  |
| variant | 0·05 (7·90[E-04] to 2·50) | 0·130 |
| variant x age^1^ | 1·17 (0·99 to 1·39) | 0·059 |
| **Any cancer females** |  |  |
| variant | 1·93 (1·38 to 2·70) | 1·12[E-04] |
| variant x age^1^ | 0·99 (0·97 to 1·01) | 0·326 |
| **Any cancer males** |  |  |
| variant | 1·83 (1·20 to 2·79) | 0·005 |
| variant x age^1^ | 0·99 (0·97 to 1·01) | 0·332 |

^1^Fit using a model of the form coxph(Surv(time,event)~variant+tt(variant)+…,tt = function(x,t, ...) x*(t-50)). The main effect is thus the estimated HR associated with carrying a variant at age 50 years. The interaction term is the estimated relative change in the HR associated with carrying a variant, per year.

***Supplemental Table 4: Cox regression analyses for the association of rMSVs in CHEK2 with cancer by type, using a continuous time-dependent coefficient***

|  | **HR (95% CI)** | ***P-*value** |
| --- | --- | --- |
| **Breast** |  |  |
| variant | 1·72 (1·22 to 2·44) | 2·21[E-03] |
| variant x age^1^ | 0·99 (0·97 to 1·01) | 0·444 |
| **Prostate** |  |  |
| variant | 1·57 (0·90 to 2·73) | 0·113 |
| variant x age^1^ | 1·00 (0·97 to 1·02) | 0·716 |
| **Pancreatic** |  |  |
| variant | 1·06 (0·19 to 6·11) | 0·946 |
| variant x age^1^ | 1·00 (0·91 to 1·09) | 0·934 |
| **Any cancer females** |  |  |
| variant | 1·22 (0·93 to 1·60) | 0·146 |
| variant x age^1^ | 1·00 (0·98 to 1·02) | 0·887 |
| **Any cancer males** |  |  |
| variant | 1·28 (0·92 to 1·79) | 0·145 |
| variant x age^1^ | 1·00 (0·98 to 1·02) | 0·914 |

^1^Fit using a model of the form coxph(Surv(time,event)~variant+tt(variant)+…,tt = function(x,t, ...) x*(t-50)). The main effect is thus the estimated HR associated with carrying a variant at age 50 years. The interaction term is the estimated relative change in the HR associated with carrying a variant, per year

| ***Supplemental Table 5a: Cox regression analyses for the association of PTVs in ATM with cancer by type using a time-dependent coefficient*** | | | | | | | | |
| --- | --- | --- | --- | --- | --- | --- | --- | --- |
|  | **All cancers** | | **Breast** | | **Prostate** | | **Pancreas** |  |
| **Age group** | **HR (95% CI)** | ***P-*value** | **HR (95% CI)** | ***P-*value** | **HR (95% CI)** | ***P-*value** | **HR (95% CI)** | ***P-*value** |
| **<50** | 4·76 (2·47 to 9·19) | 3·31[E-06] | 3·92 (2·16 to 7·10) | 6·64[E-06] | 1·98 (0·28 to 14·12) | 0·495 | 9·15 (1·26 to 66·39) | 0·029 |
| **50-59** | 3·28 (2·41 to 4·48) | 6·95[E-14] | 3·13 (2·02 to 4·87) | 3·62 [E-07] | 2·56 (1·37 to 4·76) | 3·06[E-03] | 6·24 (2·00 to 19·50) | 1·64[E-03] |
| **60-69** | 2·04 (1·60 to 2·60) | 1·05[E-08] | 1·52 (0·88 to 2·62) | 0·133 | 2·54 (1·82 to 3·54) | 3·86[E-08] | 4·91 (2·33 to 10·34) | 2·82[E-05] |
| **70-79** | 2·20 (1·74 to 2·78) | 5·02[E-11] | 1·69 (0·81 to 3·56) | 0·164 | 2·89 (1·88 to 4·43) | 1·27[E-06] | 6·48 (2·90 to 14·50) | 5·43[E-06] |
| ***Supplemental Table 5b: Cox regression analyses for the association of rMSVs in ATM with cancer by type using a time-dependent coefficient*** | | | | | | | | |
|  | **All cancers** | | **Breast** | | **Prostate** | | **Pancreas** |  |
| **Age group** | **HR (95% CI)** | ***P-*value** | **HR (95% CI)** | ***P-*value** | **HR (95% CI)** | ***P-*value** | **HR (95% CI)** | ***P-*value** |
| **<50** | 1·41 (1·01 to 1·99) | 0·046 | 1·30 (0·97 to 1·75) | 0·079 | 1·65 (0·94 to 2·88) | 0·081 | 0·63 (0·09 to 4·58) | 0·649 |
| **50-59** | 1·03 (0·89 to 1·19) | 0·720 | 1·06 (0·86 to 1·30) | 0·577 | 0·97 (0·75 to 1·25) | 0·790 | 1·09 (0·54 to 2·21) | 0·813 |
| **60-69** | 1·13 (1·04 to 1·23) | 4·22[E-03] | 1·05 (0·90 to 1·24) | 0·528 | 1·08 (0·95 to 1·22) | 0·246 | 1·62 (1·16 to 2·25) | 4·57[E-03] |
| **70-79** | 1·07 (0·99 to 1·17) | 0·104 | 1·07 (0·84 to 1·37) | 0·575 | 1·06 (0·90 to 1·25) | 0·516 | 1·52 (1·01 to 2·29 | 0·046 |

***Supplemental Table 6a: Cox regression analyses for the association of PTVs in CHEK2 with cancer by type using a time-dependent coefficient***

|  | **All cancers** | | **Breast** | | **Prostate** | | **Pancreatic** | |
| --- | --- | --- | --- | --- | --- | --- | --- | --- |
| **Age group** | **HR (95% CI)** | ***P-*value** | **HR (95% CI)** | ***P-*value** | **HR (95% CI)** | ***P-*value** | **HR (95% CI)** | ***P-*value** |
| **<50** | 1·69 (0·80 to 3·55) | 0·170 | 1·97 (1·09 to 3·56) | 0·026 | 3·07 (1·14 to 8·24) | 0·026 | 0 (-)^a^ |  |
| **50-59** | 1·61 (1·20 to 2·16) | 1·71[E-03] | 2·20 (1·52 to 3·19) | 3·34[E-05] | 2·13 (1·38 to 3·27) | 5·92[E-04] | 0 (-)^a^ |  |
| **60-69** | 1·75 (1·48 to 2·07) | 9·33[E-11] | 2·47 (1·86 to 3·29) | 4·59[E-10] | 1·99 (1·58 to 2·51) | 6·22[E-09] | 1·15 (0·43 to 3·08) | 0·779 |
| **70-79** | 1·39 (1·15 to 1·69) | 6·91[E-04] | 1·79 (1·08 to 2·98) | 0·025 | 1·73 (1·24 to 2·40) | 1·10 [E-03] | 2·23 (0·92 to 5·37) | 0·075 |
| ^a^ no carriers  ***Supplemental Table 6b: Cox regression analyses for the association of rMSVs in CHEK2 with cancer by type using a time-dependent coefficient*** | | | | | | | | |
|  | **All cancers** | | **Breast** | | **Prostate** | | **Pancreatic** | |
| **Age group** | **HR (95% CI)** | ***P-*value** | **HR (95% CI)** | ***P-*value** | **HR (95% CI)** | ***P-*value** | **HR (95% CI)** | ***P-*value** |
| **<50** | 1·02 (0·53 to 1·97) | 0·951 | 1·46 (0·92 to 2·33) | 0·109 | 0·70 (0·18 to 2·82) | 0·618 | 1·84 (0·25 to 13·35) | 0·547 |
| **50-59** | 1·33 (1·07 to 1·66) | 0·011 | 1·83 (1·40 to 2·39) | 9·41[E-06] | 1·47 (1·03 to 2·10) | 0·033 | 1·21 (0·39 to 3·79) | 0·742 |
| **60-69** | 1·22 (1·06 to 1·39) | 4·56[E-03] | 1·53 (1·21 to 1·92) | 2·90[E-04] | 1·37 (1·13 to 1·66) | 1·29[E-03] | 0·99 (0·50 to 1·99) | 0·985 |
| **70-79** | 1·22 (1·07 to 1·39) | 3·49[E-03] | 1·18 (0·80 to 1·74) | 0·400 | 1·34 (1·04 to 1·74) | 0·023 | 1·13 (0·51 to 2·53) | 0·768 |

|  | ***Supplemental Table 7a: Odds ratios and hazard ratios for the association of rMSVs in ATM with pancreatic cancer, by domain and CADD score*** | | | | | | | | | | |
| --- | --- | --- | --- | --- | --- | --- | --- | --- | --- | --- | --- |
|  | **Domain** | **Carrier** | **Non-carriers** | **OR (95% CI)** | ***P-*value** | **Carrier count** | **Non-carriers** | **HR (95% CI)** | ***P-*value** | **Pooled estimate** | ***P-*value** |
|  |  | **count** |  |  |  |  |  |  |  | **(95% CI)** |  |
|  | **Variant outside FAT and PIK domains** | 2 | 39 | 2·02 (0·49 to 8·38) | 0·332 | 32 | 852 | 1·53 (1·08 to 2·18) | 0·018 | 1·56 (1·11 to 2·20) | 0·011 |
|  | **Variant inside FAT or PIK domain and** | 1 | 40 | 3·01 (0·41 to 21·94) | 0·276 | 10 | 874 | 1·43 (0·77 to 2·66) | 0·264 | 1·53 (0·84 to 2·79) | 0·161 |
|  | **CADD score quintiles 1-4^1^** |  |  |  |  |  |  |  |  |  |  |
|  | **Variant inside FAT or PIK domain and** | 0 | 41 | 0 |  | 5 | 879 | 2·24 (0·93 to 5·39) | 0·072 | 0·15 (0·06 to 0·35) |  |
|  | **CADD score quintile 5^2^** |  |  |  |  |  |  |  |  |  |  |
|  | ^1^<=3.74 |  |  |  |  |  |  |  |  |  |  |
|  | ^2^>3.74 |  |  |  |  |  |  |  |  |  |  |
|  | ***Supplemental Table 7b: Odds ratios and hazard ratios for the association of rMSVs in ATM with breast cancer, by domain and CADD score*** | | | | | | | | | | |
|  | **Domain** | **Carrier count** | **Non-carriers** | **OR (95% CI)** | ***P-*value** | **Carrier count** | **Non-carriers** | **HR (95% CI)** | ***P-*value** | **Pooled estimate** | ***P-*value** |
|  |  |  |  |  |  |  |  |  |  | **(95% CI)** |  |
|  | **Variant outside FAT and PIK domains** | 159 | 6116 | 1·03 (0·88 to 1·21) | 0·710 | 155 | 6001 | 1·03 (0·88 to 1·21) | 0·697 | 1·03 (0·92 to 1·15) | 0·596 |
|  | **Variant inside FAT or PIK domain and** | 45 | 6230 | 0·88 (0·66 to 1·19) | 0·417 | 62 | 6094 | 1·25 (0·98 to 1·61) | 0·078 | 1·08 (0·89 to 1·31) | 0·416 |
|  | **CADD score quintiles 1-4^1^** |  |  |  |  |  |  |  |  |  |  |
|  | **Variant inside FAT or PIK domain and** | 30 | 6245 | 1·85 (1·28 to 2·68) | 1·13[E-03] | 17 | 6139 | 1·11 (0·69 to 1·78) | 0·678 | 1·52 (1·14 to 2·04) | 4·85[E-03] |
|  | **CADD score quintile 5^2^** |  |  |  |  |  |  |  |  |  |  |
| ^1^<=3.74 | |  |  |  |  |  |  |  |  |  |  |
| ^2^>3.74 | |  |  |  |  |  |  |  |  |  |  |

|  | ***Supplemental Table 7c: Odds ratios and hazard ratios for the association of rMSVs in ATM with prostate cancer, by domain and CADD score*** | | | | | | | | | | | |
| --- | --- | --- | --- | --- | --- | --- | --- | --- | --- | --- | --- | --- |
|  | **Domain** | | **Carrier**  **count** | **Non-carriers** | **OR (95% CI)** | ***P-*value** | **Carrier count** | **Non-carriers** | **HR (95% CI)** | ***P-*value** | **Pooled estimate** | ***P-*value** |
|  |  |  |  |  |  |  |  |  |  |  | **(95% CI)** |  |
|  | **Variant outside FAT and PIK domains** | | 77 | 2485 | 1·23 (0·98 to 1·55) | 0·073 | 203 | 7962 | 1·05 (0·92 to 1·21) | 0·480 | 1·10 (0·97 to 1·23) | 0·134 |
|  | **Variant inside FAT or PIK domain and** | | 23 | 2539 | 1·14 (0·75 to 1·72) | 0·550 | 76 | 8089 | 1·16 (0·92 to 1·45) | 0·208 | 1·16 (0·94 to 1·42) | 0·164 |
|  | **CADD score quintiles 1-4^1^** | |  |  |  |  |  |  |  |  |  |  |
|  | **Variant inside FAT or PIK domain and** | | 20 | 2542 | 2·99 (1·91 to 4·69) | 1·89[E-06] | 21 | 8144 | 0·98 (0·64 to 1·50) | 0·924 | 1·67 (1·23 to 2·29) | 1·20[E-03] |
|  | **CADD score quintile 5^2^** | |  |  |  |  |  |  |  |  |  |  |
|  | ^1^<=3.74 | |  |  |  |  |  |  |  |  |  |  |
|  | ^2^>3.74 | |  |  |  |  |  |  |  |  |  |  |
|  | ***Supplemental Table 7d: Odds ratios and hazard ratios for the association of rMSVs in ATM with any cancer, by domain and CADD score*** | | | | | | | | | | | |
|  | **Domain** | | **Carrier count** | **Non-carriers** | **OR (95% CI)** | ***P-*value** | **Carrier count** | **Non-carriers** | **HR (95% CI)** | ***P-*value** | **Pooled estimate** | ***P-*value** |
|  |  |  |  |  |  |  |  |  |  |  | **(95% CI)** |  |
|  | **Variant outside FAT and PIK domains** | | 508 | 18330 | 1·11 (1·01 to 1·21) | 0·030 | 971 | 36831 | 1·07 (1·01 to 1·14) | 0·035 | 1·08 (1·03 to 1·14) | 2·45[E-03] |
|  | **Variant inside FAT or PIK domain and** | | 131 | 18707 | 0·86 (0·72 to 1·02) | 0·089 | 334 | 37468 | 1·11 (1·00 to 1·23) | 0·063 | 1·02 (0·93 to 1·13) | 0·644 |
|  | **CADD score quintiles 1-4^1^** | |  |  |  |  |  |  |  |  |  |  |
|  | **Variant inside FAT or PIK domain and** | | 76 | 18762 | 1·56 (1·23 to 1·97) | 2·16[E-04] | 122 | 37680 | 1·27 (1·06 to 1·52) | 8·54[E-03] | 1·37 (1·19 to 1·58) | 1·17[E-05] |
|  | **CADD score quintile 5^2^** | |  |  |  |  |  |  |  |  |  |  |
| ^1^<=3.74 | |  |  |  |  |  |  |  |  |  |  |  |
| ^2^>3.74 | |  |  |  |  |  |  |  |  |  |  |  |

|  | ***Supplemental Table 7e: Odds ratios and hazard ratios for the association of rMSVs in ATM with any cancer in females, by domain and CADD score*** | | | | | | | | | | | |
| --- | --- | --- | --- | --- | --- | --- | --- | --- | --- | --- | --- | --- |
|  | **Domain** | | **Carrier** | **Non-carriers** | **OR (95% CI)** | ***P-*value** | **Carrier count** | **Non-carriers** | **HR (95% CI)** | ***P-*value** | **Pooled estimate** | ***P-*value** |
|  |  |  | **count** |  |  |  |  |  |  |  | **(95% CI)** |  |
|  | **Variant outside FAT and PIK domains** | | 298 | 11537 | 1·03 (0·91 to 1·16) | 0·682 | 451 | 17262 | 1·05 (0·96 to 1·15) | 0·321 | 1·04 (0·97 to 1·12) | 0·277 |
|  | **Variant inside FAT or PIK domain and** | | 84 | 11751 | 0·87 (0·70 to 1·09) | 0·216 | 160 | 17553 | 1·13 (0·97 to 1·32) | 0·126 | 1·03 (0·91 to 1·17) | 0·642 |
|  | **CADD score quintiles 1-4^1^** | |  |  |  |  |  |  |  |  |  |  |
|  | **Variant inside FAT or PIK domain and** | | 45 | 11790 | 1·47 (1·08 to 2·00) | 0·013 | 60 | 17653 | 1·35 (1·05 to 1·74) | 0·021 | 1·40 (1·15 to 1·71) | 8·74[E-04] |
|  | **CADD score quintile 5^2^** | |  |  |  |  |  |  |  |  |  |  |
|  | ^1^<=3.74 | |  |  |  |  |  |  |  |  |  |  |
|  | ^2^>3.74 | |  |  |  |  |  |  |  |  |  |  |
|  | ***Supplemental Table 7f: Odds ratios and hazard ratios for the association of rMSVs in ATM with any cancer in males, by domain and CADD score*** | | | | | | | | | | | |
|  | **Domain** | | **Carrier** | **Non-carriers** | **OR (95% CI)** | ***P-*value** | **Carrier count** | **Non-carriers** | **HR (95% CI)** | ***P-*value** | **Pooled estimate** | ***P-*value** |
|  |  |  | **count** |  |  |  |  |  |  |  | **(95% CI)** |  |
|  | **Variant outside FAT and PIK domains** | | 210 | 6793 | 1·24 (1·08 to 1·43) | 2·79 [E-03] | 520 | 19569 | 1·10 (1·00 to 1·20) | 0·041 | 1·13 (1·06 to 1·21) | 4·43[E-04] |
|  | **Variant inside FAT or PIK domain and** | | 47 | 6956 | 0·84 (0·63 to 1·12) | 0·239 | 174 | 19915 | 1·08 (0·93 to 1·26) | 0·290 | 1·02 (0·89 to 1·17) | 0·751 |
|  | **CADD score quintiles 1-4^1^** | |  |  |  |  |  |  |  |  |  |  |
|  | **Variant inside FAT or PIK domain and** | | 31 | 6972 | 1·70 (1·18 to 2·45) | 4·45[E-03] | 62 | 20027 | 1·20 (0·94 to 1·54) | 0·151 | 1·34 (1·09 to 1·65) | 6·57[E-03] |
|  | **CADD score quintile 5^2^** | |  |  |  |  |  |  |  |  |  |  |
| ^1^<=3.74 | |  |  |  |  |  |  |  |  |  |  |  |
| ^2^>3.74 | |  |  |  |  |  |  |  |  |  |  |  |

| ***Supplemental Table 8a: Odds ratios and hazard ratios for the association of rMSVs in CHEK2 with breast cancer by Helix score*** | | | | | | | | | | |
| --- | --- | --- | --- | --- | --- | --- | --- | --- | --- | --- |
| **Category** | **Carrier count** | **Non-carriers** | **OR (95% CI)** | ***P-*value** | **Carrier count** | **Non-carriers** | **HR (95% CI)** | ***P-*value** | **Pooled estimate** | ***P-*value** |
|  | **count** |  |  |  | **count** |  |  |  | **(95% CI)** |  |
| **Helix score <=0·5** | 61 | 6214 | 1·28 (0·99 to 1·66) | 0·057 | 71 | 6085 | 1·56 (1·23 to 1·97) | 2·11[E-04] | 1·42 (1·20 to 1·69) | 6·37[E-05] |
| **Helix score >0·5** | 51 | 6224 | 1·73 (1·30 to 2·30) | 1·56[E-04] | 42 | 6114 | 1·50 (1·11 to 2·04) | 8·35[E-03] | 1·62 (1·31 to 2·01) | 9·53[E-06] |
| ***Supplemental Table 8b: Odds ratios and hazard ratios for the association of rMSVs in CHEK2 with prostate cancer by Helix score*** | | | | | | | | | | |
| **Category** | **Carrier** | **Non-carriers** | **OR (95% CI)** | ***P-*value** | **Carrier count** | **Non-carriers** | **HR (95% CI)** | ***P-*value** | **Pooled estimate** | ***P-*value** |
|  | **count** |  |  |  | **count** |  |  |  | **(95% CI)** |  |
| **Helix score <=0·5** | 25 | 2537 | 1·32 (0·88 to 1·96) | 0·176 | 86 | 8079 | 1·47 (1·19 to 1·82) | 3·70[E-04] | 1·44 (1·19 to 1·74) | 1·56[E-04] |
| **Helix score >0·5** | 17 | 2545 | 1·53 (0·94 to 2·48) | 0·085 | 44 | 8121 | 1·32 (0·98 to 1·77) | 0·067 | 1·37 (1·07 to 1·77) | 0·014 |
| ***Supplemental Table 8c: Odds ratios and hazard ratios for the association of rMSVs in CHEK2 with pancreatic cancer by Helix score*** | | | | | | | | | | |
| **Category** | **Carrier** | **Non-carriers** | **OR (95% CI)** | ***P-*value** | **Carrier count** | **Non-carriers** | **HR (95% CI)** | ***P-*value** | **Pooled estimate** | ***P-*value** |
|  | **count** |  |  |  | **count** |  |  |  | **(95% CI)** |  |
| **Helix score <=0·5** | 1 | 40 | 3·26 (0·45 to 23·76) | 0·243 | 6 | 878 | 0·94 (0·42 to 2·10) | 0·882 | 1·12 (0·53 to 2·36) | 0·761 |
| **Helix score >0·5** | 0 | 41 | 0 |  | 4 | 880 | 1·07 (0·40 to 2·87) | 0·887 | 0·37 (0·14 to 1·00) |  |

| ***Supplemental Table 8d: Odds ratios and hazard ratios for the association of rMSVs in CHEK2 with any cancer in males and females by Helix score*** | | | | | | | | | | |
| --- | --- | --- | --- | --- | --- | --- | --- | --- | --- | --- |
| **Domain** | **Carrier** | **Non-carriers** | **OR (95% CI)** | ***P-*value** | **Carrier** | **Non-carriers** | **HR (95% CI)** | ***P-*value** | **Pooled estimate** | ***P-*value** |
|  | **count** |  |  |  | **count** |  |  |  | **(95% CI)** |  |
| **Helix score <=0·5** | 174 | 18664 | 1·24 (1·06 to 1·44) | 7·18[E-03] | 326 | 37476 | 1·18 (1·06 to 1·32) | 2·38[E-03] | 1·20 (1·10 to 1·32) | 1·22[E-04] |
| **Helix score >0·5** | 131 | 18707 | 1·54 (1·29 to 1·84) | 2·46[E-06] | 212 | 37590 | 1·31 (1·14 to 1·50) | 9·99[E-05] | 1·55 (1·30 to 1·84) | 1·21[E-06] |

| ***Supplemental Table 8e: Odds ratios and hazard ratios for the association of rMSVs in CHEK2 with any cancer in females by Helix score*** | | | | | | | | | | |
| --- | --- | --- | --- | --- | --- | --- | --- | --- | --- | --- |
| **Domain** | **Carrier** | **Non-carriers** | **OR (95% CI)** | ***P-*value** | **Carrier count** | **Non-carriers** | **HR (95% CI)** | ***P-*value** | **Pooled estimate** | ***P-*value** |
|  | **count** |  |  |  | **count** |  |  |  | **(95% CI)** |  |
| **Helix score <=0·5** | 112 | 11723 | 1·26 (1·04 to 1·53) | 0·021 | 152 | 17561 | 1·16 (0·99 to 1·36) | 0·075 | 1·20 (1·06 to 1·36) | 3·72[E-03] |
| **Helix score >0·5** | 88 | 11747 | 1·60 (1·28 to 1·99) | 3·29[E-05] | 104 | 17609 | 1·30 (1·07 to 1·57) | 7·94[E-03] | 1·43 (1·23 to 1·65) | 1·60[E-06] |
| ***Supplemental Table 8f: Odds ratios and hazard ratios for the association of rMSVs in CHEK2 with any cancer in males by Helix score*** | | | | | | | | | | |
| **Domain** | **Carrier** | **Non-carriers** | **OR (95% CI)** | ***P-*value** | **Carrier count** | **Non-carriers** | **HR (95% CI)** | ***P-*value** | **Pooled estimate** | ***P-*value** |
|  | **count** |  |  |  | **count** |  |  |  | **(95% CI)** |  |
| **Helix score <=0·5** | 62 | 6941 | 1·20 (0·93 to 1·55) | 0·163 | 174 | 19915 | 1·22 (1·05 to 1·41) | 9·90[E-03] | 1·22 (1·06 to 1·39) | 4·31[E-03] |
| **Helix score >0·5** | 43 | 6960 | 1·44 (1·05 to 1·96) | 0·022 | 108 | 19981 | 1·33 (1·10 to 1·60) | 3·42[E-03] | 1·35 (1·15 to 1·60) | 3·61[E-04] |
