## Supplemental Figures for "Protein-truncating and rare missense variants in *ATM* and *CHEK2* and associations with cancer in UK Biobank whole-exome sequenced data"

Supplemental Figure 1: Cumulative absolute female breast cancer risks in *ATM* and *CHEK2* protein-truncating/missense variant carriers and the general female population by age

Supplemental Figure 2: Cumulative absolute prostate cancer risks in *ATM* and *CHEK2* protein-truncating/missense variant carriers and the general male population by age

Supplemental Figure 3: Cumulative absolute any cancer (other than non melanoma skin cancer) risks for female *ATM* and *CHEK2* protein-truncating/missense variant carriers and the general female population by age

Supplemental Figure 4: Cumulative absolute any cancer (other than non melanoma skin cancer) risks for male *ATM* and *CHEK2* protein-truncating/missense variant carriers and the general male population by age

Supplemental Figure 1: Cumulative absolute female breast cancer1 risks in ATM and CHEK2 protein-truncating/missense variant carriers and the general female population by age

Cumulative risk

| Age | ATM PTV | ATM MSV | CHEK2 PTV | CHEK2 MSV | General population risk |
| --- | --- | --- | --- | --- | --- |
| 20 | 0.000105557 | 5.52881e-06 | 1.11781e-05 | 7.60139e-06 | 4.99999e-06 |
| 21 | 0.00039663 | 2.32208e-05 | 4.69472e-05 | 3.19254e-05 | 2.09998e-05 |
| 22 | 0.000673698 | 4.09125e-05 | 8.2715e-05 | 5.62489e-05 | 3.69993e-05 |
| 23 | 0.000937409 | 5.86039e-05 | 0.000118482 | 8.05718e-05 | 5.29986e-05 |
| 24 | 0.00118839 | 7.62949e-05 | 0.000154247 | 0.000104894 | 6.89976e-05 |
| 25 | 0.00142722 | 9.39857e-05 | 0.000190011 | 0.000129216 | 8.49964e-05 |
| 26 | 0.00305953 | 0.000221129 | 0.000447027 | 0.000304011 | 0.00019998 |
| 27 | 0.00461055 | 0.000348256 | 0.000703977 | 0.000478775 | 0.00031495 |
| 28 | 0.00608431 | 0.000475366 | 0.000960862 | 0.000653509 | 0.000429908 |
| 29 | 0.00748463 | 0.000602461 | 0.00121768 | 0.000828212 | 0.000544852 |
| 30 | 0.00881517 | 0.000729539 | 0.00147443 | 0.00100288 | 0.000659782 |
| 31 | 0.0122413 | 0.00107423 | 0.00217068 | 0.00147663 | 0.000971528 |
| 32 | 0.0154898 | 0.0014188 | 0.00286645 | 0.00195014 | 0.00128318 |
| 33 | 0.0185702 | 0.00176325 | 0.00356173 | 0.00242344 | 0.00159473 |
| 34 | 0.0214913 | 0.00210758 | 0.00425653 | 0.00289651 | 0.00190618 |
| 35 | 0.0242617 | 0.00245179 | 0.00495085 | 0.00336935 | 0.00221754 |
| 36 | 0.0297951 | 0.00317734 | 0.00641356 | 0.00436584 | 0.00287386 |
| 37 | 0.0350287 | 0.00390236 | 0.00787413 | 0.00536133 | 0.00352976 |
| 38 | 0.0399797 | 0.00462685 | 0.00933256 | 0.00635583 | 0.00418522 |
| 39 | 0.0446644 | 0.00535081 | 0.0107889 | 0.00734933 | 0.00484025 |
| 40 | 0.0490978 | 0.00607425 | 0.012243 | 0.00834184 | 0.00549485 |
| 41 | 0.0570286 | 0.00744273 | 0.0149908 | 0.0102186 | 0.00673323 |
| 42 | 0.0645082 | 0.00880932 | 0.017731 | 0.0120918 | 0.00797007 |
| 43 | 0.0715647 | 0.010174 | 0.0204635 | 0.0139614 | 0.00920537 |
| 44 | 0.0782243 | 0.0115369 | 0.0231885 | 0.0158275 | 0.0104391 |
| 45 | 0.0845112 | 0.0128978 | 0.025906 | 0.0176901 | 0.0116714 |
| 46 | 0.0947219 | 0.0152396 | 0.0305729 | 0.0208928 | 0.013792 |
| 47 | 0.104323 | 0.0175758 | 0.0352176 | 0.0240851 | 0.0159081 |
| 48 | 0.113356 | 0.0199065 | 0.0398401 | 0.027267 | 0.0180197 |
| 49 | 0.121857 | 0.0222317 | 0.0444405 | 0.0304385 | 0.0201267 |
| 50 | 0.129863 | 0.0245513 | 0.049019 | 0.0335997 | 0.0222292 |
| 51 | 0.139673 | 0.0275646 | 0.0549501 | 0.0377021 | 0.0249612 |
| 52 | 0.148895 | 0.0305687 | 0.0608444 | 0.0417871 | 0.0276855 |
| 53 | 0.157569 | 0.0335634 | 0.066702 | 0.0458549 | 0.0304023 |
| 54 | 0.16573 | 0.0365489 | 0.0725233 | 0.0499053 | 0.0331114 |
| 55 | 0.173414 | 0.0395252 | 0.0783083 | 0.0539387 | 0.035813 |
| 56 | 0.180797 | 0.0425527 | 0.0841741 | 0.0580366 | 0.0385618 |
| 57 | 0.187752 | 0.0455706 | 0.0900027 | 0.0621167 | 0.0413028 |
| 58 | 0.194307 | 0.048579 | 0.0957944 | 0.0661793 | 0.044036 |
| 59 | 0.200488 | 0.0515779 | 0.101549 | 0.0702242 | 0.0467614 |
| 60 | 0.206317 | 0.0545673 | 0.107268 | 0.0742517 | 0.049479 |
| 61 | 0.212822 | 0.0580933 | 0.113989 | 0.0789959 | 0.0526854 |
| 62 | 0.218954 | 0.0616062 | 0.120659 | 0.0837159 | 0.0558809 |
| 63 | 0.224736 | 0.0651059 | 0.12728 | 0.0884117 | 0.0590657 |
| 64 | 0.230191 | 0.0685926 | 0.133851 | 0.0930834 | 0.0622398 |
| 65 | 0.235339 | 0.0720663 | 0.140372 | 0.0977313 | 0.0654031 |
| 66 | 0.241265 | 0.0762873 | 0.148264 | 0.10337 | 0.0692485 |
| 67 | 0.246853 | 0.0804891 | 0.156083 | 0.108974 | 0.0730781 |
| 68 | 0.252124 | 0.0846718 | 0.16383 | 0.114543 | 0.076892 |
| 69 | 0.257098 | 0.0888355 | 0.171507 | 0.120078 | 0.0806901 |
| 70 | 0.261794 | 0.0929802 | 0.179113 | 0.125577 | 0.0844726 |
| 71 | 0.265804 | 0.0967107 | 0.185929 | 0.130519 | 0.0878784 |
| 72 | 0.269593 | 0.100426 | 0.192688 | 0.135433 | 0.0912716 |
| 73 | 0.273175 | 0.104126 | 0.199392 | 0.14032 | 0.0946521 |
| 74 | 0.276563 | 0.10781 | 0.20604 | 0.145178 | 0.0980201 |
| 75 | 0.279766 | 0.11148 | 0.212633 | 0.15001 | 0.101375 |
| 76 | 0.283043 | 0.11543 | 0.219701 | 0.155203 | 0.10499 |
| 77 | 0.286142 | 0.119364 | 0.226705 | 0.160365 | 0.108589 |
| 78 | 0.289074 | 0.12328 | 0.233646 | 0.165496 | 0.112174 |
| 79 | 0.291934 | 0.127178 | 0.240526 | 0.170595 | 0.115745 |
| 80 | 0.294781 | 0.131059 | 0.247344 | 0.175663 | 0.119302 |
| 81 | 0.29781 | 0.135185 | 0.254558 | 0.181041 | 0.123084 |
| 82 | 0.300826 | 0.139291 | 0.261703 | 0.186385 | 0.12685 |
| 83 | 0.303829 | 0.143378 | 0.26878 | 0.191693 | 0.1306 |
| 84 | 0.306819 | 0.147445 | 0.275789 | 0.196967 | 0.134334 |
| 85 | 0.309796 | 0.151493 | 0.282732 | 0.202207 | 0.138052 |

^1^For breast cancer in *ATM* truncating variant carriers, where there was evidence of a decline of risk with age, HRs from the time-dependent model in Supplemental Table 1 were used. In all other cases a constant HR was assumed.

Supplmental Figure 2: Cumulative absolute prostate cancer risks for CHEK2 and ATM protein truncating/missense carriers and the general male population by age

Cumulative risk

| Age | ATM PTV | ATM MSV | CHEK2 PTV | CHEK2 MSV | General population Risk |
| --- | --- | --- | --- | --- | --- |
| 10 | 0 | 0 | 0 | 0 | 0 |
| 11 | 0 | 0 | 0 | 0 | 0 |
| 12 | 0 | 0 | 0 | 0 | 0 |
| 13 | 0 | 0 | 0 | 0 | 0 |
| 14 | 0 | 0 | 0 | 0 | 0 |
| 15 | 0 | 0 | 1.94954e-06 | 0 | 0 |
| 16 | 2.55099e-06 | 1.07704e-06 | 3.89908e-06 | 1.41307e-06 | 9.99999e-07 |
| 17 | 5.10197e-06 | 2.15408e-06 | 5.84862e-06 | 2.82614e-06 | 2e-06 |
| 18 | 7.65294e-06 | 3.23112e-06 | 7.79815e-06 | 4.2392e-06 | 3e-06 |
| 19 | 1.02039e-05 | 4.30815e-06 | 9.74768e-06 | 5.65227e-06 | 3.99999e-06 |
| 20 | 1.27549e-05 | 5.38519e-06 | 9.74768e-06 | 7.06533e-06 | 4.99999e-06 |
| 21 | 1.27549e-05 | 5.38519e-06 | 9.74768e-06 | 7.06533e-06 | 4.99999e-06 |
| 22 | 1.27549e-05 | 5.38519e-06 | 9.74768e-06 | 7.06533e-06 | 4.99999e-06 |
| 23 | 1.27549e-05 | 5.38519e-06 | 9.74768e-06 | 7.06533e-06 | 4.99999e-06 |
| 24 | 1.27549e-05 | 5.38519e-06 | 9.74768e-06 | 7.06533e-06 | 4.99999e-06 |
| 25 | 1.27549e-05 | 5.38519e-06 | 9.74768e-06 | 7.06533e-06 | 4.99999e-06 |
| 26 | 1.27549e-05 | 5.38519e-06 | 9.74768e-06 | 7.06533e-06 | 4.99999e-06 |
| 27 | 1.27549e-05 | 5.38519e-06 | 9.74768e-06 | 7.06533e-06 | 4.99999e-06 |
| 28 | 1.27549e-05 | 5.38519e-06 | 9.74768e-06 | 7.06533e-06 | 4.99999e-06 |
| 29 | 1.27549e-05 | 5.38519e-06 | 9.74768e-06 | 7.06533e-06 | 4.99999e-06 |
| 30 | 1.27549e-05 | 5.38519e-06 | 1.16972e-05 | 7.06533e-06 | 4.99999e-06 |
| 31 | 1.53058e-05 | 6.46222e-06 | 1.36467e-05 | 8.47839e-06 | 5.99998e-06 |
| 32 | 1.78568e-05 | 7.53926e-06 | 1.55962e-05 | 9.89144e-06 | 6.99998e-06 |
| 33 | 2.04077e-05 | 8.61629e-06 | 1.75457e-05 | 1.13045e-05 | 7.99997e-06 |
| 34 | 2.29586e-05 | 9.69332e-06 | 1.94953e-05 | 1.27176e-05 | 8.99996e-06 |
| 35 | 2.55096e-05 | 1.07703e-05 | 2.72933e-05 | 1.41306e-05 | 9.99995e-06 |
| 36 | 3.57132e-05 | 1.50785e-05 | 3.50912e-05 | 1.97828e-05 | 1.39999e-05 |
| 37 | 4.59168e-05 | 1.93865e-05 | 4.28891e-05 | 2.54349e-05 | 1.79998e-05 |
| 38 | 5.61202e-05 | 2.36946e-05 | 5.06869e-05 | 3.10871e-05 | 2.19998e-05 |
| 39 | 6.63235e-05 | 2.80027e-05 | 5.84846e-05 | 3.67392e-05 | 2.59997e-05 |
| 40 | 7.65268e-05 | 3.23107e-05 | 0.000142307 | 4.23912e-05 | 2.99996e-05 |
| 41 | 0.000186205 | 7.86209e-05 | 0.000226122 | 0.000103149 | 7.29973e-05 |
| 42 | 0.000295871 | 0.000124929 | 0.00030993 | 0.000163903 | 0.000115993 |
| 43 | 0.000405525 | 0.000171235 | 0.000393731 | 0.000224653 | 0.000158987 |
| 44 | 0.000515167 | 0.000217539 | 0.000477525 | 0.0002854 | 0.00020198 |
| 45 | 0.000624797 | 0.00026384 | 0.000876911 | 0.000346142 | 0.00024497 |
| 46 | 0.00114729 | 0.000484551 | 0.00127614 | 0.00063568 | 0.000449899 |
| 47 | 0.00166951 | 0.000705213 | 0.00167521 | 0.000925133 | 0.000654786 |
| 48 | 0.00219145 | 0.000925826 | 0.00207412 | 0.0012145 | 0.00085963 |
| 49 | 0.00271312 | 0.00114639 | 0.00247287 | 0.00150379 | 0.00106443 |
| 50 | 0.00323452 | 0.00136691 | 0.00394395 | 0.00179299 | 0.00126919 |
| 51 | 0.00515753 | 0.00218078 | 0.00541286 | 0.0028602 | 0.00202495 |
| 52 | 0.00707684 | 0.00299399 | 0.00687962 | 0.00392627 | 0.00278013 |
| 53 | 0.00899245 | 0.00380654 | 0.00834421 | 0.0049912 | 0.00353474 |
| 54 | 0.0109044 | 0.00461843 | 0.00980666 | 0.00605499 | 0.00428878 |
| 55 | 0.0128126 | 0.00542965 | 0.0136947 | 0.00711765 | 0.00504224 |
| 56 | 0.0178816 | 0.00758897 | 0.0175675 | 0.00994493 | 0.00704805 |
| 57 | 0.0229247 | 0.00974361 | 0.0214252 | 0.0127642 | 0.0090498 |
| 58 | 0.0279419 | 0.0118936 | 0.0252677 | 0.0155754 | 0.0110475 |
| 59 | 0.0329334 | 0.0140389 | 0.0290952 | 0.0183786 | 0.0130412 |
| 60 | 0.0378993 | 0.0161795 | 0.0358127 | 0.0211739 | 0.0150309 |
| 61 | 0.0466002 | 0.0199456 | 0.0424839 | 0.026087 | 0.0185321 |
| 62 | 0.0552226 | 0.0236972 | 0.049109 | 0.0309756 | 0.0220209 |
| 63 | 0.0637672 | 0.0274345 | 0.0556884 | 0.0358396 | 0.0254973 |
| 64 | 0.0722347 | 0.0311575 | 0.0622223 | 0.0406793 | 0.0289613 |
| 65 | 0.0806257 | 0.0348663 | 0.0735398 | 0.0454947 | 0.0324131 |
| 66 | 0.0951171 | 0.0413176 | 0.084721 | 0.0538573 | 0.0384195 |
| 67 | 0.109381 | 0.0477257 | 0.0957676 | 0.0621467 | 0.0443887 |
| 68 | 0.12342 | 0.0540911 | 0.106681 | 0.0703636 | 0.0503208 |
| 69 | 0.137238 | 0.0604139 | 0.117463 | 0.0785086 | 0.056216 |
| 70 | 0.150839 | 0.0666944 | 0.130444 | 0.0865823 | 0.0620747 |
| 71 | 0.167144 | 0.0743009 | 0.143234 | 0.096338 | 0.0691741 |
| 72 | 0.183137 | 0.0818454 | 0.155836 | 0.10599 | 0.0762197 |
| 73 | 0.198824 | 0.0893285 | 0.168253 | 0.115538 | 0.083212 |
| 74 | 0.21421 | 0.0967506 | 0.180489 | 0.124985 | 0.0901513 |
| 75 | 0.229301 | 0.104112 | 0.194234 | 0.134331 | 0.0970382 |
| 76 | 0.246172 | 0.112441 | 0.207749 | 0.144877 | 0.104835 |
| 77 | 0.262674 | 0.120693 | 0.221038 | 0.155294 | 0.112564 |
| 78 | 0.278816 | 0.128867 | 0.234105 | 0.165584 | 0.120227 |
| 79 | 0.294605 | 0.136966 | 0.246953 | 0.175749 | 0.127823 |
| 80 | 0.310049 | 0.14499 | 0.257219 | 0.185791 | 0.135354 |
| 81 | 0.322331 | 0.151445 | 0.267345 | 0.193848 | 0.141416 |
| 82 | 0.334394 | 0.157851 | 0.277334 | 0.201826 | 0.147436 |
| 83 | 0.346244 | 0.164209 | 0.287187 | 0.209725 | 0.153414 |
| 84 | 0.357882 | 0.170519 | 0.296906 | 0.217546 | 0.159349 |
| 85 | 0.369314 | 0.176781 |  | 0.225289 | 0.165243 |

Supplemental Figure 3: Cumulative absolute any cancer (other than non melanoma skin cancer)1 risks for female CHEK2 and ATM protein truncating/missense variant carriers and the general female population by age

Cumulative risk

| Age | ATM PTV | ATM Missense | CHEK2 PTV | CHEK2 MSV | General population risk |
| --- | --- | --- | --- | --- | --- |
| 0 | 0 | 0 | 0 | 0 | 0 |
| 1 | 0 | 0 | 0 | 0 | 0.000186983 |
| 2 | 0.00279824 | 0.000203173 | 0.00031117 | 0.000223835 | 0.00037393 |
| 3 | 0.00550831 | 0.000406304 | 0.000622243 | 0.00044762 | 0.000560843 |
| 4 | 0.00813308 | 0.000609394 | 0.000933219 | 0.000671355 | 0.00074772 |
| 5 | 0.0106753 | 0.000812443 | 0.0012441 | 0.00089504 | 0.000934563 |
| 6 | 0.0120192 | 0.00092318 | 0.00141363 | 0.00101703 | 0.00103646 |
| 7 | 0.0133224 | 0.0010339 | 0.00158313 | 0.001139 | 0.00113835 |
| 8 | 0.0145861 | 0.00114462 | 0.0017526 | 0.00126096 | 0.00124023 |
| 9 | 0.0158115 | 0.00125532 | 0.00192205 | 0.00138291 | 0.0013421 |
| 10 | 0.0169997 | 0.001366 | 0.00209146 | 0.00150484 | 0.00144396 |
| 11 | 0.0182875 | 0.0014897 | 0.00228077 | 0.00164109 | 0.00155779 |
| 12 | 0.0195361 | 0.00161338 | 0.00247005 | 0.00177733 | 0.0016716 |
| 13 | 0.0207468 | 0.00173704 | 0.00265929 | 0.00191355 | 0.0017854 |
| 14 | 0.0219206 | 0.00186069 | 0.0028485 | 0.00204975 | 0.00189919 |
| 15 | 0.0230588 | 0.00198433 | 0.00303766 | 0.00218593 | 0.00201297 |
| 16 | 0.0249942 | 0.00220119 | 0.00336945 | 0.0024248 | 0.00221255 |
| 17 | 0.0268693 | 0.00241801 | 0.00370113 | 0.00266362 | 0.00241209 |
| 18 | 0.028686 | 0.00263478 | 0.0040327 | 0.00290237 | 0.00261158 |
| 19 | 0.0304462 | 0.0028515 | 0.00436416 | 0.00314107 | 0.00281104 |
| 20 | 0.0321517 | 0.00306818 | 0.0046955 | 0.00337971 | 0.00301046 |
| 21 | 0.0349017 | 0.00342884 | 0.00524695 | 0.00377693 | 0.0033424 |
| 22 | 0.0375635 | 0.00378937 | 0.0057981 | 0.00417398 | 0.00367423 |
| 23 | 0.0401401 | 0.00414977 | 0.00634894 | 0.00457088 | 0.00400595 |
| 24 | 0.0426343 | 0.00451004 | 0.00689947 | 0.00496761 | 0.00433757 |
| 25 | 0.0450489 | 0.00487018 | 0.0074497 | 0.00536419 | 0.00466907 |
| 26 | 0.0501032 | 0.00564949 | 0.00864 | 0.00622231 | 0.00538644 |
| 27 | 0.0549833 | 0.0064282 | 0.00982887 | 0.00707969 | 0.0061033 |
| 28 | 0.0596956 | 0.0072063 | 0.0110163 | 0.00793633 | 0.00681964 |
| 29 | 0.0642466 | 0.00798378 | 0.0122023 | 0.00879224 | 0.00753546 |
| 30 | 0.0686423 | 0.00876066 | 0.0133869 | 0.0096474 | 0.00825077 |
| 31 | 0.0755525 | 0.0100254 | 0.0153144 | 0.0110395 | 0.00941539 |
| 32 | 0.0822098 | 0.0112886 | 0.0172382 | 0.0124296 | 0.0105786 |
| 33 | 0.0886247 | 0.0125501 | 0.0191581 | 0.0138177 | 0.0117405 |
| 34 | 0.0948073 | 0.01381 | 0.0210744 | 0.0152039 | 0.0129011 |
| 35 | 0.100767 | 0.0150683 | 0.0229868 | 0.0165882 | 0.0140602 |
| 36 | 0.109171 | 0.0169085 | 0.0257813 | 0.0186122 | 0.0157556 |
| 37 | 0.11725 | 0.0187452 | 0.0285678 | 0.020632 | 0.017448 |
| 38 | 0.125021 | 0.0205785 | 0.0313463 | 0.0226477 | 0.0191375 |
| 39 | 0.132497 | 0.0224083 | 0.0341168 | 0.0246592 | 0.0208241 |
| 40 | 0.139689 | 0.0242347 | 0.0368795 | 0.0266665 | 0.0225078 |
| 41 | 0.150258 | 0.0270224 | 0.0410909 | 0.0297297 | 0.0250782 |
| 42 | 0.160389 | 0.0298022 | 0.0452839 | 0.0327832 | 0.0276418 |
| 43 | 0.170104 | 0.032574 | 0.0494586 | 0.0358271 | 0.0301986 |
| 44 | 0.179424 | 0.0353378 | 0.053615 | 0.0388614 | 0.0327488 |
| 45 | 0.188367 | 0.0380938 | 0.0577534 | 0.0418862 | 0.0352922 |
| 46 | 0.201827 | 0.0424123 | 0.0642253 | 0.0466242 | 0.0392788 |
| 47 | 0.214674 | 0.0467114 | 0.0706528 | 0.0513387 | 0.0432489 |
| 48 | 0.226942 | 0.0509913 | 0.0770362 | 0.0560299 | 0.0472026 |
| 49 | 0.238662 | 0.0552519 | 0.0833758 | 0.0606979 | 0.05114 |
| 50 | 0.249863 | 0.0594934 | 0.089672 | 0.0653428 | 0.0550611 |
| 51 | 0.264707 | 0.0653564 | 0.0983504 | 0.0717601 | 0.0604836 |
| 52 | 0.278829 | 0.071183 | 0.106946 | 0.0781333 | 0.065875 |
| 53 | 0.292272 | 0.0769732 | 0.11546 | 0.0844628 | 0.0712354 |
| 54 | 0.305075 | 0.0827273 | 0.123893 | 0.0907488 | 0.0765651 |
| 55 | 0.317275 | 0.0884455 | 0.132246 | 0.0969917 | 0.0818642 |
| 56 | 0.332078 | 0.095688 | 0.142785 | 0.104893 | 0.0885796 |
| 57 | 0.346132 | 0.102873 | 0.153196 | 0.112725 | 0.0952458 |
| 58 | 0.359486 | 0.110001 | 0.163481 | 0.120489 | 0.101863 |
| 59 | 0.37218 | 0.117072 | 0.173642 | 0.128184 | 0.108432 |
| 60 | 0.384255 | 0.124087 | 0.183679 | 0.135813 | 0.114953 |
| 61 | 0.399522 | 0.133348 | 0.196864 | 0.145874 | 0.123569 |
| 62 | 0.413972 | 0.142511 | 0.209837 | 0.155818 | 0.1321 |
| 63 | 0.427658 | 0.151578 | 0.2226 | 0.165647 | 0.140549 |
| 64 | 0.44063 | 0.160548 | 0.235158 | 0.175361 | 0.148915 |
| 65 | 0.452934 | 0.169424 | 0.247513 | 0.184962 | 0.157199 |
| 66 | 0.468215 | 0.180936 | 0.263433 | 0.197399 | 0.167955 |
| 67 | 0.482632 | 0.192288 | 0.279017 | 0.209646 | 0.178574 |
| 68 | 0.496244 | 0.203483 | 0.294271 | 0.221706 | 0.189057 |
| 69 | 0.509108 | 0.214523 | 0.309203 | 0.233583 | 0.199406 |
| 70 | 0.521273 | 0.225409 | 0.32382 | 0.245278 | 0.209623 |
| 71 | 0.534882 | 0.238113 | 0.340738 | 0.258903 | 0.221559 |
| 72 | 0.547715 | 0.250607 | 0.357232 | 0.272283 | 0.233315 |
| 73 | 0.559826 | 0.262897 | 0.373315 | 0.285421 | 0.244893 |
| 74 | 0.571264 | 0.274986 | 0.388996 | 0.298322 | 0.256297 |
| 75 | 0.582076 | 0.286876 | 0.404284 | 0.31099 | 0.267528 |
| 76 | 0.594525 | 0.301124 | 0.422427 | 0.326142 | 0.281006 |
| 77 | 0.606247 | 0.315088 | 0.440018 | 0.34096 | 0.294236 |
| 78 | 0.617295 | 0.328772 | 0.457073 | 0.355453 | 0.307222 |
| 79 | 0.627715 | 0.342183 | 0.47361 | 0.369627 | 0.31997 |
| 80 | 0.637552 | 0.355326 | 0.489643 | 0.38349 | 0.332483 |
| 81 | 0.647944 | 0.369733 | 0.507019 | 0.398652 | 0.346222 |
| 82 | 0.65774 | 0.383817 | 0.523804 | 0.413442 | 0.359678 |
| 83 | 0.666982 | 0.397587 | 0.540018 | 0.427867 | 0.372857 |
| 84 | 0.675709 | 0.411049 | 0.555681 | 0.441939 | 0.385765 |
| 85 | 0.683955 | 0.424211 | 0.570811 | 0.455664 | 0.398408 |

^1^For any cancer in females in *ATM* truncating variant carriers, where there was evidence of a decline of risk with age, HRs from the time-dependent model in Supplemental Table 1 were used. In all other cases a constant HR was assumed.

Supplemental Figure 4: Cumulative absolute any cancer (other than non melanoma skin cancer) risks for male CHEK2 and ATM male protein truncating/missense variant carriers and the general male population by age

Cumulative risk

| Age | ATM PTV | ATM MSV | CHEK2 PTV | CHEK2 MSV | General population risk |
| --- | --- | --- | --- | --- | --- |
| 10 | 0.00330464 | 0.00153465 | 0.00209345 | 0.00175837 | 0.000114 |
| 11 | 0.00360837 | 0.00167583 | 0.00228599 | 0.00192012 | 0.000114 |
| 12 | 0.00391201 | 0.001817 | 0.00247849 | 0.00208184 | 0.000114 |
| 13 | 0.00421556 | 0.00195815 | 0.00267096 | 0.00224354 | 0.000114 |
| 14 | 0.00451902 | 0.00209928 | 0.00286339 | 0.00240521 | 0.000114 |
| 15 | 0.00482238 | 0.00224038 | 0.00305578 | 0.00256686 | 0.0002 |
| 16 | 0.00527373 | 0.00245037 | 0.00334206 | 0.0028074 | 0.0004 |
| 17 | 0.00572487 | 0.00266031 | 0.00362825 | 0.00304788 | 0.0006 |
| 18 | 0.00617581 | 0.0028702 | 0.00391437 | 0.00328831 | 0.0008 |
| 19 | 0.00662654 | 0.00308005 | 0.0042004 | 0.00352867 | 0.001 |
| 20 | 0.00707707 | 0.00328986 | 0.00448636 | 0.00376898 | 0.001333 |
| 21 | 0.00777593 | 0.00361541 | 0.00493001 | 0.00414184 | 0.001666 |
| 22 | 0.00847429 | 0.00394086 | 0.00537347 | 0.00451457 | 0.001999 |
| 23 | 0.00917217 | 0.00426619 | 0.00581674 | 0.00488715 | 0.002332 |
| 24 | 0.00986955 | 0.00459142 | 0.0062598 | 0.0052596 | 0.002665 |
| 25 | 0.0105664 | 0.00491655 | 0.00670267 | 0.00563191 | 0.003386 |
| 26 | 0.0116506 | 0.00542257 | 0.00739185 | 0.00621133 | 0.004107 |
| 27 | 0.0127335 | 0.00592834 | 0.00808054 | 0.00679041 | 0.004828 |
| 28 | 0.0138153 | 0.00643385 | 0.00876876 | 0.00736916 | 0.005549 |
| 29 | 0.0148958 | 0.0069391 | 0.0094565 | 0.00794757 | 0.00627 |
| 30 | 0.0159752 | 0.0074441 | 0.0101438 | 0.00852564 | 0.007445 |
| 31 | 0.017548 | 0.00818046 | 0.0111457 | 0.00936847 | 0.00862 |
| 32 | 0.0191182 | 0.00891626 | 0.0121466 | 0.0102106 | 0.009795 |
| 33 | 0.020686 | 0.00965153 | 0.0131464 | 0.011052 | 0.01097 |
| 34 | 0.0222512 | 0.0103862 | 0.0141453 | 0.0118927 | 0.012145 |
| 35 | 0.023814 | 0.0111204 | 0.0151432 | 0.0127327 | 0.013866 |
| 36 | 0.0258439 | 0.012075 | 0.0164402 | 0.0138247 | 0.015587 |
| 37 | 0.0278696 | 0.0130287 | 0.0177355 | 0.0149155 | 0.017308 |
| 38 | 0.0298911 | 0.0139814 | 0.0190291 | 0.0160051 | 0.019029 |
| 39 | 0.0319084 | 0.0149332 | 0.020321 | 0.0170935 | 0.02075 |
| 40 | 0.0339215 | 0.0158841 | 0.0216112 | 0.0181807 | 0.023383 |
| 41 | 0.0367479 | 0.017221 | 0.0234243 | 0.0197088 | 0.026016 |
| 42 | 0.039566 | 0.018556 | 0.0252341 | 0.0212346 | 0.028649 |
| 43 | 0.0423758 | 0.0198892 | 0.0270404 | 0.0227581 | 0.031282 |
| 44 | 0.0451775 | 0.0212206 | 0.0288435 | 0.0242791 | 0.033915 |
| 45 | 0.047971 | 0.0225502 | 0.0306432 | 0.0257978 | 0.038056 |
| 46 | 0.0527863 | 0.024847 | 0.0337499 | 0.0284206 | 0.042197 |
| 47 | 0.0575772 | 0.0271384 | 0.0368468 | 0.0310362 | 0.046338 |
| 48 | 0.062344 | 0.0294244 | 0.0399337 | 0.0336449 | 0.050479 |
| 49 | 0.0670867 | 0.031705 | 0.0430107 | 0.0362465 | 0.05462 |
| 50 | 0.0718054 | 0.0339803 | 0.0460779 | 0.0388411 | 0.060375 |
| 51 | 0.0803895 | 0.0381353 | 0.0516722 | 0.043577 | 0.06613 |
| 52 | 0.0888944 | 0.0422724 | 0.0572337 | 0.0482896 | 0.071885 |
| 53 | 0.0973208 | 0.0463918 | 0.0627627 | 0.0529789 | 0.07764 |
| 54 | 0.105669 | 0.0504934 | 0.0682593 | 0.0576452 | 0.083395 |
| 55 | 0.113941 | 0.0545774 | 0.0737237 | 0.0622885 | 0.090736 |
| 56 | 0.128708 | 0.0619198 | 0.0835261 | 0.070629 | 0.098077 |
| 57 | 0.14323 | 0.0692052 | 0.0932249 | 0.0788954 | 0.105418 |
| 58 | 0.15751 | 0.076434 | 0.102821 | 0.0870883 | 0.112759 |
| 59 | 0.171552 | 0.0836067 | 0.112316 | 0.0952084 | 0.1201 |
| 60 | 0.185361 | 0.0907237 | 0.12171 | 0.103256 | 0.129882 |
| 61 | 0.207313 | 0.102172 | 0.136766 | 0.116183 | 0.139664 |
| 62 | 0.228675 | 0.113476 | 0.151564 | 0.128923 | 0.149446 |
| 63 | 0.249462 | 0.124639 | 0.166108 | 0.14148 | 0.159228 |
| 64 | 0.26969 | 0.13566 | 0.180404 | 0.153856 | 0.16901 |
| 65 | 0.289373 | 0.146543 | 0.194454 | 0.166054 | 0.181854 |
| 66 | 0.318573 | 0.162989 | 0.215563 | 0.184444 | 0.194698 |
| 67 | 0.346574 | 0.179118 | 0.23612 | 0.202428 | 0.207542 |
| 68 | 0.373427 | 0.194937 | 0.256138 | 0.220017 | 0.220386 |
| 69 | 0.399178 | 0.21045 | 0.275633 | 0.237217 | 0.23323 |
| 70 | 0.423871 | 0.225665 | 0.294616 | 0.254039 | 0.248447 |
| 71 | 0.454733 | 0.245181 | 0.318767 | 0.275544 | 0.263664 |
| 72 | 0.483944 | 0.264204 | 0.342092 | 0.29643 | 0.278881 |
| 73 | 0.511592 | 0.282748 | 0.364619 | 0.316714 | 0.294098 |
| 74 | 0.537761 | 0.300825 | 0.386375 | 0.336414 | 0.309315 |
| 75 | 0.562529 | 0.318447 | 0.407386 | 0.355546 | 0.327887 |
| 76 | 0.591689 | 0.339893 | 0.432693 | 0.378733 | 0.346459 |
| 77 | 0.618909 | 0.360665 | 0.45692 | 0.401087 | 0.365031 |
| 78 | 0.644315 | 0.380784 | 0.480112 | 0.422637 | 0.383603 |
| 79 | 0.668029 | 0.40027 | 0.502316 | 0.443412 | 0.402175 |
| 80 | 0.690164 | 0.419142 | 0.523571 | 0.46344 | 0.422972 |
| 81 | 0.712299 | 0.438753 | 0.545393 | 0.484151 | 0.443769 |
| 82 | 0.732855 | 0.457702 | 0.566216 | 0.504063 | 0.464566 |
| 83 | 0.751943 | 0.476012 | 0.586086 | 0.523207 | 0.485363 |
| 84 | 0.769668 | 0.493703 | 0.605046 | 0.541612 | 0.50616 |
| 85 | 0.786127 | 0.510797 | 0.623139 | 0.559307 | 0.528389 |
